# Pathophysiology of voluntary motor commands at the level of the spinal motoneuron in patients with multiple sclerosis

**DOI:** 10.1101/2025.08.12.25333527

**Authors:** Laura M. McPherson, Keith R. Lohse, Skyler M. Simon, James A. Beauchamp, Francesco Negro, Robert T. Naismith, Anne H. Cross

## Abstract

Multiple sclerosis (MS) is a progressive inflammatory neurodegenerative disease that degrades neural transmission between the brain and spinal α-motoneurons. These voluntary motor commands contain excitatory, inhibitory, and neuromodulatory components that must be appropriately balanced for skilled motor control. Unlike other clinical populations, in MS we have no knowledge about how voluntary motor commands are disrupted. MS is a clinically heterogeneous population, with sensorimotor impairments that vary widely and unpredictably across patients. Our overall scientific hypothesis is that the voluntary motor command in the MS population varies accordingly, with multiple “phenotypes” evident. Here, we explore this idea by identifying pathological aspects of the voluntary motor command in 59 participants with MS with a range of sensorimotor symptoms and disability, compared with 38 age-/sex-matched controls. We recorded motor unit discharge from the tibialis anterior and soleus muscles during isometric dorsiflexion/plantarflexion contractions. We then calculated geometric and temporal features in their firing patterns to characterize their excitatory, inhibitory, and neuromodulatory inputs according to a recently developed “reverse engineering” paradigm. MS values for many of our parameters were highly variable, with some participants with abnormally values and others with abnormally low values. In addition to this variability, MS group means for most parameters reflecting the balance of neuromodulation and inhibition were significantly lower than those of controls. These initial findings support the idea that there may be different phenotypes of voluntary motor command pathology among patients with MS, indicating the potential need to personalize the selection of mechanistically targeted rehabilitation therapies.

## 1. Introduction

Multiple sclerosis is an immune-mediated disease of the central nervous system (CNS) that results in demyelination and neuroaxonal degeneration that can degrade neural control of nearly all major physiological systems (Compston & Coles, 2008; Oh, 2022). The motor system is commonly affected, with up to 84% of patients experiencing a decline in motor function (Swingler, 1992; Rizzo *et al*., 2004; Johansson *et al*., 2007). Unlike hemiparetic stroke and spinal cord injury populations where the range of typical motor impairments is comparatively limited and relatively predictable, motor impairments in MS vary widely and unpredictably across patients (Oh, 2022). They commonly include neurally-mediated muscle weakness, hypo- or hypertonia, spasticity, spasms, balance deficits, discoordination, and/or ataxia (Younger, 2023).

Neurological deficits in MS emerge in two ways: episodic relapses (sudden new or worsening neurological symptoms driven by focal inflammatory demyelination (McGinley *et al*., 2021)) and disease progression (a gradual advancement of symptoms associated with “smoldering” neurodegenerative processes that occur diffusely throughout the brain and spinal cord (Giovannoni *et al*., 2022)). Both relapse-associated and progressive symptoms lead to accumulating disability over time (Kappos *et al*., 2020; Lublin *et al*., 2022), often due to motor deficits that degrade gait and balance (LaRocca, 2011; Wagner *et al*., 2014; Younger, 2023).

Little is known about the neuropathophysiological mechanisms that underlie potential motor impairments in MS, thereby limiting the development of novel, mechanistically based therapies that are emerging in other populations with neurological injury (Ellis *et al*., 2009; Thompson & Wolpaw, 2015; McPherson *et al*., 2015, 2018b; Christiansen *et al*., 2018; Jo *et al*., 2023). In part, this is because MS is so heterogeneous in terms of lesion locations, clinical symptoms, and disease course. This heterogeneity is a challenge for conventional neurophysiological approaches used in motor control research that yield few outcome measures, rely on group analyses, and/or focus primarily on the brain without considering the substantial processing of motor commands that occurs in the spinal cord.

Here, we employ a novel paradigm for studying pathological neural control of movement in MS: characterization of voluntary motor commands at the level of the spinal α-motoneuron. This approach utilizes reverse engineering of human motor unit discharge patterns to characterize the three canonical types of neural inputs to motoneurons: excitatory, inhibitory, and neuromodulatory (Johnson *et al*., 2017; Beauchamp *et al*., 2023b; Chardon *et al*., 2023). Focus on these inputs has provided insights otherwise inaccessible in other populations with neurological injury (Gorassini *et al*., 2004; McPherson *et al*., 2018c, 2018b; Trajano *et al*., 2023; Beauchamp *et al*., 2023a; Mahrous *et al*., 2024) and has motivated entirely new lines of research for developing targeted pharmacologic and rehabilitation therapies (Murray *et al*., 2011b; McPherson *et al*., 2018a, 2018b; Rich *et al*., 2020). The rich multi-dimensional information it provides could be particularly powerful in MS because it has the potential to explicate the motor heterogeneity unique to this population.

Voluntary motor commands originate cortically and are shaped at all levels of the neuraxis, culminating as excitatory, inhibitory, and neuromodulatory inputs to α-motoneurons in the spinal cord. Excitatory and inhibitory inputs convey specific task-related commands to motoneurons (Heckman & Enoka, 2012), and neuromodulatory inputs dramatically transform motoneuron excitability to meet task demands, amplifying a motoneuron’s response to excitatory and inhibitory inputs (Heckman *et al*., 2009; Heckman & Enoka, 2012). Excitation and neuromodulation are both required for strong and/or sustained muscle contractions, and inhibition is a crucial counterpart for each – synergizing with excitation to increase the complexity of motor commands and counterbalancing neuromodulation to grade muscle contractions and prevent hyperexcitability (Johnson & Heckman, 2010; Johnson *et al*., 2017).

Excitatory, inhibitory, and neuromodulatory components of the motor command must be appropriately balanced to produce skilled motor control (Heckman *et al*., 2009; Johnson *et al*., 2017). A disruption of this balance, as can occur with CNS damage, has deleterious effects on motor control, evidenced by work in the spinal cord injury (Gorassini *et al*., 2004; Mahrous *et al*., 2024), hemiparetic stroke (Mottram *et al*., 2014; McPherson *et al*., 2018c, 2018b; Beauchamp *et al*., 2023a, 2024a), and amyotrophic lateral sclerosis (Trajano *et al*., 2023) populations. Even more mild changes within the CNS, such as those associated with aging, are reflected in measurements of the voluntary motor command (Hassan *et al*., 2021; Orssatto *et al*., 2021).

Evidence to date indicates that each of these populations may have distinct neurophysiological phenotypes of voluntary motor command pathology, presumably depending on the ways in which the CNS is affected. For example, while motoneuron hyperexcitability at rest is a classic feature of people with hemiparetic stroke, recent findings indicate that pathologically high *inhibitory* input to motoneurons during voluntary contractions may be a key contributor to weakness (Mottram *et al*., 2014; McPherson *et al*., 2018c; Beauchamp *et al*., 2023a, 2024a). For people with spinal cord injury, loss of neuromodulatory input causes profound weakness acutely after injury, which gradually improves when motoneuron receptors that would normally receive neuromodulatory input undergo compensatory adaptations (Murray *et al*., 2010; Rank *et al*., 2011).

Because patients with MS have multiple lesions that can be located anywhere throughout the CNS, our overall scientific hypothesis is that multiple neurophysiological phenotypes of voluntary motor command pathology are evident in the MS population. Here, we explore this idea by identifying and characterizing pathological aspects of the voluntary motor command in a heterogeneous sample of patients with MS by reverse engineering motor unit discharge (Johnson *et al*., 2017; Chardon *et al*., 2023) from the tibialis anterior and soleus muscles.

## 2. Methods

### 2.1. Participants

Participants with MS were recruited from the John L. Trotter Multiple Sclerosis Center at Washington University and/or the Washington University Physical Therapy outpatient clinic. Individuals without neurological injury or disease (controls) were recruited through the Washington University Research Participant Registry or by word of mouth. They were included in the study if their sex and age (within 5 years) matched those of an MS participant.

Potential participants were screened prior to study enrollment. Participants with MS were required to have (1) a confirmed diagnosis of MS made by a neurologist, (2) the ability to generate a muscle contraction in at least one of the tested muscles for ≥ 20 sec, (3) had no clinical relapses within the last six months, (4) the ability to transfer into the testing setup with physical therapist assistance if needed, and (5) sufficient passive range of motion in the tested leg to sit comfortably in the testing setup.

Participants from both groups were excluded if they (1) had a history of severe musculoskeletal or peripheral neurological injury to the tested leg, (2) pain or low skin integrity that would prevent sitting in the testing chair, (3) severe cognitive or visual deficits, (4) uncontrolled hypertension or (5) central neurological injury or disease (other than MS for the MS group).

Fifty-nine individuals with MS met all enrollment criteria and completed the study. Data from 58 participants were included in the analysis (47 females, 11 males; mean age ± SD: 51.3 ± 10.8 years, range 29-79; mean symptom duration ± SD: 16.6 ± 10.3 years, range: <1 - 44; Table 1), because no useable motor units could be obtained from one participant. Participant disability was assessed using the Expanded Disease Disability Scale (EDSS), and enrolled participants had scores ranging from 0 (no disability) to 7 (severe disability). The mean ± SD time to complete a forward 25-ft walk was 7.8 ± 7.3 sec (range 3.72 – 54.0), and the mean ± SD time to complete a backward 25-ft walk (Edwards *et al*., 2020) was 16.2 ± 19.0 sec (range 3.52 – 122.2).

**Table 1.** Demographics and disease information for the participants with MS.

|  | Age range | Sex | Disease subtype | Years since symptom onset | EDSS | S1 DTR |
| --- | --- | --- | --- | --- | --- | --- |
| S01 | 30-34 | F | RRMS | 0-4 | 0 | 2 |
| S02 | 50-54 | F | RRMS | 35-39 | 3 | 0 |
| S03 | 60-64 | F | SPMS | 35-39 | 3 | 2 |
| S04 | 50-54 | F | RRMS | 15-19 | 3.5 | 4 |
| S05 | 45-50 | F | SPMS | 5-9 | 3.5 | 3 |
| S06 | 25-29 | F | RRMS | 5-9 | 0 | 2 |
| S07 | 50-55 | F | RRMS | 10-14 | 1.5 | 2 |
| S08 | 45-49 | F | SPMS | 20-24 | 6 | 4 (patellar) |
| S09 | 50-54 | F | RRMS | 20-24 | 1.5 | 2 |
| S10 | 60-64 | F | PPMS | 10-14 | 4 | 3 |
| S11 | 60-64 | M | RRMS | 20-24 | 1.5 | 2 (patellar) |
| S12 | 65-69 | F | RRMS | 20-24 | 3.5 | 2 |
| S13 | 65-69 | F | PPMS | 15-19 | 5 | 2 |
| S14 | 50-54 | F | RRMS | 20-24 | 3.5 | 1 |
| S15 | 40-44 | F | RRMS | 5-9 | 1.5 | 2 |
| S16 | 60-64 | M | PPMS | 10-14 | 6 | 1 (patellar) |
| S17 | 65-69 | F | RRMS | 35-39 | 4 | 0 |
| S18 | 35-39 | M | RRMS | 10-14 | 2 | 2 |
| S19 | 70-74 | F | PPMS | 20-24 | 6 | 2 |
| S20 | 45-49 | F | RRMS | 25-29 | 3 | 3 |
| S21 | 35-39 | F | SPMS | 15-19 | 6.5 | 4 (patellar) |
| S22 | 45-49 | M | RRMS | 10-14 | 3 | 2 |
| S23 | 55-59 | F | SPMS | 20-24 | 4.5 | 3 |
| S24 | 60-64 | F | RRMS | 25-29 | 2 | 2 |
| S25 | 50-54 | F | RRMS | 0-4 | 2 | 2 |
| S26 | 60-64 | F | RRMS | 25-29 | 2 | 2 |
| S27 | 55-59 | F | PPMS | 10-14 | 5.5 | 2 |
| S28 | 75-79 | F | SPMS | 40-44 | 5.5 | 0 |
| S29 | 45-49 | F | RRMS | 10-14 | 1 | 2 |
| S30 | 40-44 | F | RRMS | 0-4 | 1 | 2 |
| S31 | 45-49 | F | RRMS | 10-14 | 1 | 2 |
| S32 | 40-44 | F | RRMS | 5-9 | 0 | 2 |
| S33 | 35-39 | M | RRMS | 10-14 | 0 | 1 (patellar) |
| S34 | 60-64 | F | SPMS | 25-29 | 5.5 | 4 |
| S35 | 45-49 | F | RRMS | 15-19 | 2 | 2 |
| S36 | 40-44 | F | RRMS | 0-4 | 0 | 2 |
| S37 | 50-54 | F | RRMS | 0-4 | 4 | 3 |
| S38 | 40-44 | M | RRMS | 0-4 | 1 | 2 |
| S39 | 65-69 | F | RRMS | 30-34 | 3.5 | 2 (patellar) |
| S40 | 60-64 | F | RRMS | 25-29 | 2 | 2 |
| S41 | 60-64 | F | SPMS | 5-9 | 3 | 2 |
| S42 | 55-59 | F | SPMS | 30-34 | 7 | 2 |
| S43 | 50-55 | F | RRMS | 30-34 | 3 | 3-4 (1 beat clonus) |
| S44 | 40-44 | F | SPMS | 5-9 | 3 | 4 (4 beats clonus) |
| S45 | 60-64 | F | SPMS | 15-19 | 1.5 | 3 |
| S46 | 45-49 | M | RRMS | 0-4 | 2 | 2 |
| S47 | 40-44 | M | RRMS | 5-9 | 3 | 2 |
| S48 | 40-44 | F | RRMS | 0-4 | 3 | 2 |
| S49 | 50-54 | F | RRMS | 15-19 | 2 | 2 |
| S50 | 40-44 | M | SPMS | 10-14 | 7 | 5 (18 beats clonus) |
| S51 | 30-34 | F | RRMS | 5-9 | 2 | 2 |
| S52 | 45-49 | F | RRMS | 15-19 | 2 | 2 |
| S53 | 45-49 | F | RRMS | 25-29 | 0 | 2 (patellar) |
| S54 | 45-49 | M | RRMS | 25-29 | 2 | 3 (patellar) |
| S55 | 65-69 | M | PPMS | 5-9 | 6 | 0 |
| S56 | 45-49 | F | RRMS | 10-14 | 2.5 | 1 |
| S57 | 35-39 | F | RRMS | 5-9 | 1.5 | 2 |
| S58 | 55-59 | F | SRMS | 5-9 | 5 | 2 |
| Mean<br>± SD | 51.2 ± 10.9 | 46<br>F/<br>11<br>M | 40 RRMS<br>13 SPMS<br>5 PPMS | 16.6 ± 10.4 | 3.0 ± 1.9 |  |

**EDSS: Expanded Disability Status Scale.** (0: no disability; 1.0-4.5: can walk without an assistive device, increasing levels of impairment in any of eight functional systems of the body; 5.0: walking limited to 200m; 5.5: walking limited to 100m; 6.0: walking requires unilateral assistive device; 6.5: walking up to 20 m requires bilateral support; 7.0: Unable to walk > 5m; able to propel self in manual wheelchair. Max score: 10 (death due to MS.). **Deep tendon reflex (DTR) score.** 0: absent reflex; 1: trace response or only seen with reinforcement; 2: normal; 3: brisk; 4: brisk with non-sustained clonus; 5: brisk with sustained clonus. When S1 DTR scores were not available, L2-L4 (patellar) DTRs are presented.

Thirty-eight individuals without neurological injury met all enrollment criteria and completed the study (26 females, 12 males; mean age ± SD: 47.6 ± 15.6 years, range 24-76). For each participant with MS, there was an average of four controls in the dataset who were of matched age and sex (range 1-9). All participants provided written informed consent for participation in the study, which was approved by the Institutional Review Board of Washington University.

### 2.2. Experimental Setup and Data Collection

Experimental testing was conducted using the Biodex System 4 Pro dynamometer (Biodex Medical Systems, Shirley, NY) (**Figure 1**) to measure isometric joint torque. Participants sat upright in the experimental chair with their tested foot secured to the ankle attachment of the dynamometer. Their leg was positioned at 90° hip flexion and 10° ankle plantarflexion, which resulted in approximately 20° knee flexion. A computer monitor displayed real-time visual feedback of torque data.

**Figure 1.**
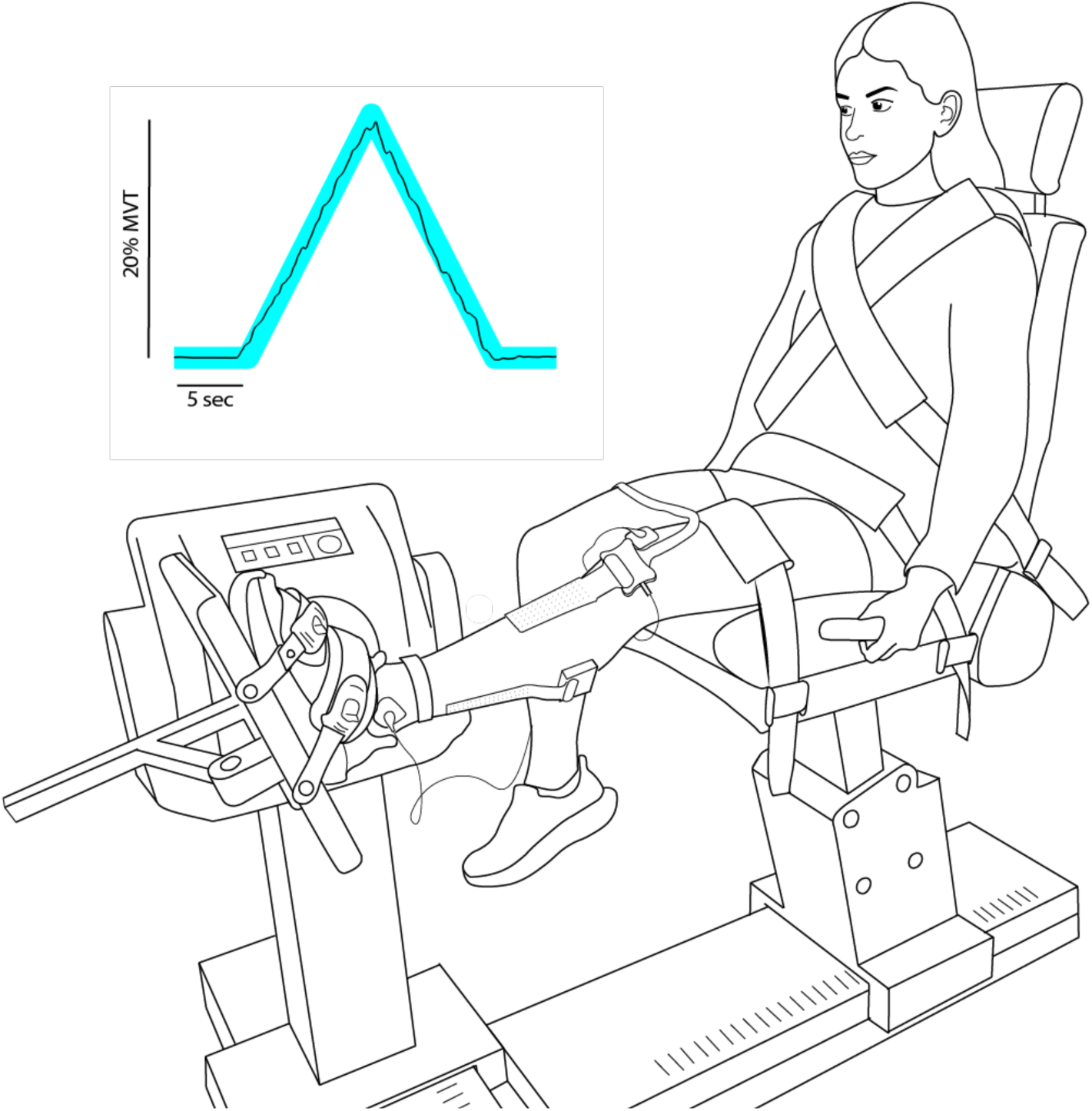
Experimental setup. Participants were seated with the tested foot secured to the dynamometer. Real-time visual feedback of torque generation during experimental trials (inset) was displayed on a TV monitor.

High-density surface EMG was recorded in monopolar fashion from 64-channel rectangular electrode grids with 8 mm interelectrode distance (GR08MM1305, OT Bioelettronica, Inc.) secured to skin that had been prepared using abrasive paste. One electrode grid was placed over the tibialis anterior muscle belly, and two electrode grids were placed over the soleus muscle belly, one on each side of the Achilles tendon. Care was taken to ensure the electrode remained on the soleus and did not cover the lateral ankle musculature. Analog EMG and torque signals were acquired using the Quattrocento data acquisition system (OT Bioelettronica, Inc.) with a sampling rate of 2048 Hz, high-pass filter of 10 Hz, and low-pass filter of 900 Hz.

### 2.3. Experimental Protocol

Participants completed the protocol with both legs, performed in randomized order. In this analysis, we include data from the MS participants’ more impaired leg and from the control participants’ dominant leg. The MS participants’ more impaired leg was determined based on patient report of motor symptoms. For the participants who did not have appreciable motor symptoms, the more impaired leg was determined based on sidedness of non-motor symptoms or chosen randomly.

First, each participant’s isometric maximum voluntary torques (MVTs) were measured in dorsiflexion and plantarflexion, performed in randomized order. Trials within a direction were repeated until three trials within 85% of the maximum value were obtained, without the last one being the highest. Participants were given vigorous verbal encouragement during MVT trials and several minutes of rest in between trials.

Then, participants performed submaximal triangular contractions with a visual template provided on the screen (blue line in Figure 1). They were instructed to rest for 5 sec at trial initiation, then slowly increase their isometric torque at a rate of 2% MVT/sec over 10 sec to a peak of 20% MVT, and then slowly decrease their isometric torque at the same rate prior to resting for another 5 sec. Trials were repeated until the participant completed three to five trials with appropriately smooth and triangular torque traces (typically 5-10 total per direction). The triangular contractions were performed in both dorsiflexion and plantarflexion, with the trial blocks for each direction completed in randomized order.

### 2.4. Data Analysis

#### 2.4.1. Motor unit decomposition and processing

Offline, high-density surface EMG channels were visually inspected, and channels with substantial noise and/or movement artifacts were removed from further analysis. The remaining channels were decomposed automatically into individual motor unit spike trains using a convolutive blind source separation algorithm with a silhouette threshold of 0.85 (Negro *et al*., 2016). The accuracy of each resulting spike train was inspected by a trained team member and improved iteratively using a well-validated local re-optimization technique implemented using a custom MATLAB graphical user interface, to correct minor errors from the automatic decomposition or discard the motor unit (Del Vecchio *et al*., 2020; Hug *et al*., 2021; Hassan *et al*., 2021; Jenz *et al*., 2023). Motor units were tracked across trials using cross-correlation of their motor unit action potential profiles (Martinez-Valdes *et al*., 2017; Thompson *et al*., 2018).

A custom MATLAB program was used to define a motor unit’s recruitment time as the first spike time to be followed by sustained firing with subsequent inter-spike intervals with duration less than 500 ms. A motor unit’s de-recruitment time was defined as the last spike time before an inter-spike interval of greater than 500 ms.

We used a support vector machine regression model with kernel value of 2.2 to predict smoothed motor unit discharge profiles from instantaneous discharge rates with accurate onset and offset discharge rates and low levels of residual error (Beauchamp *et al*., 2022). Torque signals were smoothed using an acausal one-sided moving average filter with window length of 250 ms, baseline corrected, and normalized to the MVT value for that torque direction.

#### 2.4.2. Reverse engineering parameters

Reverse engineering parameters are calculated from the trajectory of motor unit discharge rate over time during the standardized slowly increasing and decreasing triangle contraction. The premise is that the shape of this trajectory will change according to the mixture of excitatory, inhibitory, and neuromodulatory input to the motoneuron. Based on several decades of animal work and simulations using realistic motoneuron models, various geometric and temporal parameters of the discharge rate trajectory have been identified that provide information about the three types of inputs (Johnson *et al*., 2017; Beauchamp *et al*., 2022; Chardon *et al*., 2023; McPherson *et al*., 2024).

Understanding our reverse engineering parameters and interpreting our results in subsequent sections first requires a brief discussion about how neuromodulatory and inhibitory inputs control motoneuron excitability through facilitation and deactivation of persistent inward currents (PICs) in motoneuron dendrites. Neuromodulatory input is delivered via the raphespinal and ceruleospinal tracts which descend from brainstem nuclei, project diffusely to multiple motor pools, and release serotonin and norepinephrine, respectively (Holstege & Kuypers, 1987). These neurotransmitters profoundly increase motoneuron excitability by facilitating the activation of PICs, which are long lasting depolarizing currents generated by voltage-sensitive Na^+^ and Ca^2+^ channels that change how the motoneuron processes and responds to excitatory and inhibitory inputs (i.e., its input/output function) (Heckman *et al*., 2005, 2009; Heckman & Enoka, 2012). The amplitude of PICs and the associated change in motoneuron excitability is proportional to the amount of neuromodulatory input to motoneuron dendrites (Heckman *et al*., 2005).

The effect of PICs on a motoneuron’s input/output function happens in three distinctive ways (amplification of excitatory input, saturation of responsiveness to excitatory input, prolongation of excitatory input) that consequently induce three corresponding non-linearities in motoneuron firing patterns (acceleration, attenuation, recruitment hysteresis). Figure 2A depicts how these distinctive non-linearities, which are readily observed in human motor unit firing patterns, are magnified with higher PIC amplitude.

**Figure 2.**
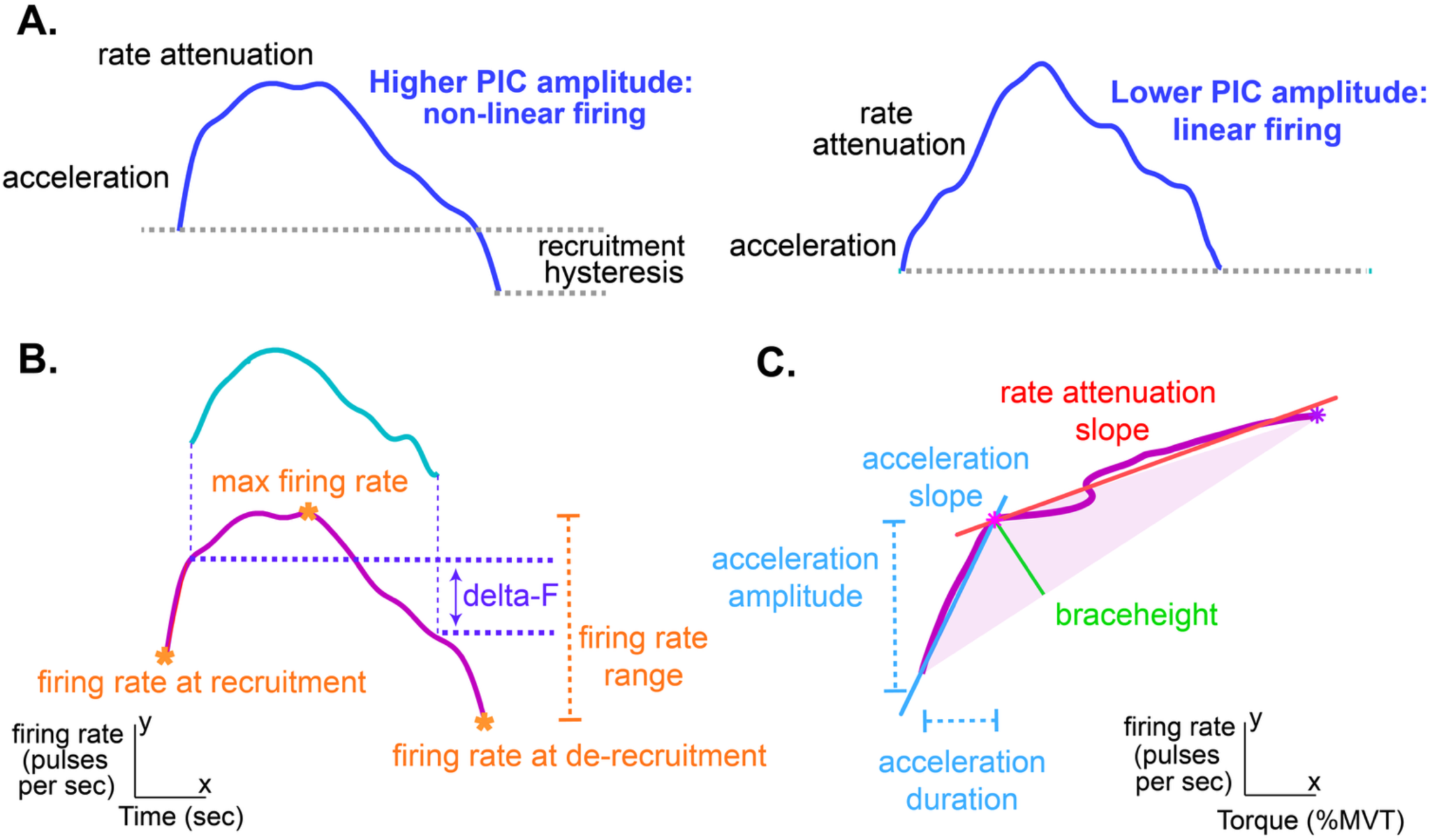
A. The effect of PIC amplitude on a motor unit’s firing pattern. Left: during a triangular contraction, motor units with large PICs produce firing patterns with a characteristic non-linear shape. Three distinct phases can be seen: rapid initial acceleration, rate attenuation marked by a more gradual rate increase, and recruitment hysteresis where the motor unit de-recruits at a lower level of excitatory synaptic input than it was recruited. The example shown is a motor unit from the TA of a control participant. Right: Motor units with small (or no) PICs produce more linear firing patterns. **B. Calculation of reverse engineering parameters from the firing rate vs. time function.** Calculation of firing rate parameters (maximum, range, value at recruitment, value at de-recruitment) and delta-F are shown. Additional variables calculated from the firing rate vs. time function include normalized delta-F, duration ratio, torque at recruitment and derecruitment, duration, and recruitment range. **C. Calculation of reverse engineering parameters from the firing rate vs. torque function.** Calculation of acceleration parameters (amplitude, slope, duration), rate attenuation slope, and braceheight are shown.

In contrast to neuromodulatory input, inhibitory synaptic input is highly effective at *reducing* PIC amplitude. It also interacts with excitatory input and changes the net synaptic excitatory drive. Thus, its effects are also apparent in human motor unit firing patterns. Inhibitory input can occur in patterns that are either reciprocal or proportional to excitatory input, which have dramatically different effects on PIC amplitude and motor unit firing patterns (Powers *et al*., 2012; Powers & Heckman, 2017; Johnson *et al*., 2017; Chardon *et al*., 2023), discussed in more detail below.

##### Reverse engineering variables quantifying firing rate acceleration and attenuation

As the PIC is activated near a motoneuron’s spiking threshold, it amplifies the depolarizing current delivered by excitatory synaptic input by up to five-fold, enabling the motoneuron to fire repetitively and reach an optimal firing rate (Binder *et al*., 2020). We quantify the magnitude of this rapid PIC activation in motor unit firing patterns as the initial acceleration slope, amplitude, and duration (**Figure 2A, 2C**). Subsequently, when dendrites have strong PIC depolarization, they become “saturated” to further increases in excitatory input as the excitatory driving force into the motoneuron is reduced, resulting in an attenuation of the speed of firing rate increase (**Figure 2A, 2C**).

The pattern of inhibition has a substantial effect on the rate attenuation slope (Powers & Heckman, 2017; Beauchamp *et al*., 2023b). Proportional “balanced” inhibition (i.e., inhibition that increases proportionally to excitation) reduces the net excitation and dramatically lowers the rate attenuation slope and overall rate modulation; it also lowers PIC amplitude. With reciprocal “push-pull” inhibition (i.e., inhibition that decreases proportionally to increases in excitation), PICs are high in amplitude but are activated gradually, which reduces initial acceleration and imparts a steep positive rate attenuation slope.

We calculate acceleration and attenuation parameters on the smoothed firing rate vs. torque function as follows (**Figure 2C**). First, we quantify the overall non-linearity of increasing motor unit discharge by calculating the extent to which the firing rate trajectory from recruitment to maximal firing rate deviates from a theoretical linear increase. This parameter, braceheight, is calculated as the magnitude of the maximal orthogonal distance between the firing rate trajectory and a straight line between recruitment and maximal firing rate (**Figure 2C**), then normalized to the maximum possible braceheight value that would be obtained from a theoretical right triangle with the same start and end points (Beauchamp *et al*., 2023b). The location of the maximal orthogonal distance on the firing rate vs. torque function is designated as the point of inflection separating the acceleration and attenuation phases. Acceleration slope (pps/%MVT) is calculated as the change in firing rate during the acceleration phase divided by the corresponding change in torque. Acceleration amplitude and duration are the amplitude (pps) and duration (%MVT) of the acceleration phase. Rate attenuation slope (pps/%MVT) is calculated as the linear regression slope for the segment of the firing rate vs. torque function between the inflection point and the maximal firing rate (Beauchamp *et al*., 2023b). Some motor units peak quickly after the acceleration phase, complicating calculation of the rate attenuation slope. Therefore, if the duration of the rate attenuation phase was less than 1 second, we calculated the rate attenuation slope over the firing rate vs. torque function from the inflection point to 1 second prior to the maximal torque. This allowed for calculation of negative rate attenuation slopes when found.

Each of these parameters reflect aspects of PIC amplification and saturation and thus could theoretically be sensitive to changes in both neuromodulation level and the pattern of inhibition. However, recent studies using realistic motoneuron simulations indicated that each parameter has a different sensitivity to these two types of inputs (Beauchamp *et al*., 2023b; Chardon *et al*., 2023). Based on these findings, our current interpretation is that acceleration slope is sensitive to both neuromodulation and inhibition, rate attenuation slope is relatively specific to inhibition, and braceheight is relatively specific to neuromodulation.

##### Reverse engineering variables quantifying recruitment hysteresis

The final characteristic of PICs, that they prolong the effect of excitatory synaptic input, results in self-sustained motoneuron firing that persists even after excitatory synaptic input decreases below the level required for recruitment. This effect is evident in human motor unit firing patterns as recruitment-de-recruitment hysteresis, where the motor unit is de-recruited at a lower contraction level (i.e., level of excitatory drive) than it was recruited (**Figure 2A, 2B**).

Here, we quantify recruitment-derecruitment hysteresis using the “delta-F” paired motor unit technique (Gorassini *et al*., 2002), which is the most established method for estimating PIC amplitude in humans (Mesquita *et al*., 2024) and has been validated in animal models and using motoneuron simulations (Powers & Heckman, 2015). Motor units within a trial are compared in a pairwise fashion, calculating delta-F for the higher threshold motor unit as the difference in firing rate of the lower threshold motor unit between the times of recruitment and de-recruitment of the higher threshold motor unit (**Figure 2B**). We used the following criteria to identify appropriate motor unit pairs (Gorassini *et al*., 2002; Hassan *et al*., 2021; Jenz *et al*., 2023): 1) recruitment time difference of > 1 sec, 2) correlation of the smoothed discharge rates with R^2^ > 0.7, and 3) rate modulation of the control unit that is at least 0.5 pps greater than the delta-F value.

One critique of the delta-F technique is that the level of rate modulation in the low threshold motor unit may partially confound the measure. For example, delta-F calculated with a low threshold motor unit with minimal rate modulation could underestimate PIC amplitude. Thus, we also used a normalized delta-F, which is the calculated delta-F divided by the total possible delta-F (using the lower threshold motor unit’s minimum firing rate during the descending portion of the contraction rather than the firing rate at the time of high threshold de-recruitment). For both forms of delta-F, we obtained one value per motor unit by averaging over all values calculated with different lower threshold motor units (Hassan *et al*., 2021; Jenz *et al*., 2023).

Finally, we calculated each motor unit’s self-sustained firing duration ratio to estimate hysteresis in time by calculating the firing duration on the descending contraction minus the firing duration on the ascending contraction, divided by the total firing duration (Hassan *et al*., 2021; Mohammadalinejad *et al*., 2024).

Based on the motoneuron simulations discussed above, delta-F is sensitive to changes in both neuromodulation level and the pattern of inhibition (Beauchamp *et al*., 2023b; Chardon *et al*., 2023). The self-sustained duration ratio has not been included in simulations yet, but we interpret it the same as delta-F.

##### Reverse engineering variables related to recruitment, firing rate, and de-recruitment

For each motor unit recording, we calculated the following parameters from the smoothed discharge profile: firing rate at recruitment and de-recruitment, maximal firing rate, firing rate range (maximum firing rate minus minimum firing rate), and total firing duration. We calculated torque at the times of recruitment and de-recruitment for each motor unit, and the recruitment range for each trial (the torque at recruitment of the highest threshold motor unit minus the torque at recruitment of the lowest threshold motor unit).

Firing rate parameters (recruitment, de-recruitment, maximal, range) are affected by both neuromodulatory and inhibitory inputs (Wienecke *et al*., 2009; Powers *et al*., 2012; Powers & Heckman, 2017), in addition to motoneuron afterhyperpolarization duration (dependent on input conductance that varies with motoneuron size) (Manuel *et al*., 2005).

Parameters related to recruitment and de-recruitment also provide information about the distribution of excitatory input on low vs. high threshold motor units (Johnson *et al*., 2017; Chardon *et al*., 2023), which differs based on the source of excitatory input (e.g., corticospinal vs. Ia afferent) and impacts when motoneurons are recruited and de-recruited during an increasing contraction. Based on recent reverse engineering simulations (Chardon *et al*., 2023), torque at de-recruitment, recruitment range, total firing duration, and torque at recruitment provide the most information about the distribution of excitatory input.

### 2.5. Statistical Analyses

All statistical analyses were performed in R (version 4.4.0, (R Core Team, 2024)) with statistical significance set to ⍺ = 0.05. As described below, we used different linear mixed-effect regression models to address different research questions (Bates *et al*., 2022). Each of these models included specific fixed effects (depending on the tested hypotheses) and random effects (to account for statistical dependencies in how the data were structured).

#### 2.5.1. Group differences in maximal strength

We compared maximal strength between groups using a linear mixed effects model with fixed effects of *Group*, *Torque Direction*, and *Group*-by-*Torque Direction*, along with covariates of *Age_c* (grand mean-centered *Age*) and *Sex* and a random intercept of *Participant*. *Group, Torque Direction*, and *Sex* were contrast coded so that the associated regression coefficients would correspond to the mean difference between the two levels of each factor (as: MS - Control, dorsiflexion - plantarflexion, Male - Female).

#### 2.5.2. Group differences in motor unit parameters

To assess between-group differences for each motor unit parameter, we performed a linear mixed effects model with fixed effects of *Group*, *Muscle*, *Group-by-Muscle*, and covariates of *Sex* and *Age_c* as participant-level predictors. Torque at recruitment was grand-mean centered (*TQ_recrt_c*) and included as a fixed effect, serving as a motor unit-level covariate (except for the model with *TQ_recrt* as the outcome measure) (Jenz *et al*., 2023).

To fully account for the dependencies within the data, we included random intercepts for *Participant, Trial,* and *Motor Unit* within *Muscle* within *Participant* as well as random slopes for *Muscle* within *Participant* and *Muscle* within *Trial* (R syntax: (1+Muscle | Participant) + (1+Muscle | Trial) + (1 | Participant : MotorUnit : Muscle)). For some outcome variables, the random slope of *Muscle* within *Trial* did not account for any estimated variance and was therefore removed from the model to avoid a singularity. For a subset of these variables, the same scenario occurred with the random intercept of *Trial. Group, Muscle*, and *Sex* were contrast coded so that the associated coefficients would correspond to the mean difference between the two levels of each factor (coded as: MS - Control, TA - soleus, Male - Female). The coefficient of the *Group* x *Muscle* interaction is interpreted as the mean difference between groups of the mean difference between muscles. Mean differences and coefficients are reported along with their 95% confidence intervals.

Visual inspection of Q-Q plots for many of the linear mixed effects models revealed moderate-to-large departures from normality. Therefore, we applied parametric bootstrapping methods to all models using the *pbmodcomp* function from the *pbkrtest* package (Halekoh & Højsgaard, 2014) with 2000 simulations. We used the Bartlett statistic to determine statistical significance of fixed effects (Halekoh & Højsgaard, 2014).

#### 2.5.3. Heterogeneity and distributional differences between groups

For the following analyses, we aggregated values for each reverse engineering variable across motor unit recordings per participant. First, for each motor unit that was identified in multiple trials, we averaged its values across trials to provide one value per variable per motor unit. Then, we calculated the median value across all motor units, resulting in one value per variable per person.

Because MS group presents itself in the clinic in such a qualitatively different way from other clinical populations, we chose a variety of approaches to try to capture the nature of the MS population. To compare how the MS and control samples were distributed around their means, we examined characteristics of the shapes of the distributions. First, we tested for statistical differences between the MS and control distribution shapes for each parameter by centering the distributions around their group mean and applying the two-tailed two-sample Kolmogorov-Smirnov test (*stats* package included in R). Then, we calculated the interquartile range, range, and coefficient of variation of the non-mean-centered distributions for each parameter for each muscle. Given that the duration ratio means were near zero, we reported the variance instead of the coefficient of variation to avoid inflated values. We tested for equality of variance between groups per parameter and muscle using Levene’s test from the *car* package (Fox & Weisberg, 2019). For the MS and control distributions, there were occasionally distributions with an extreme value that was substantially separate from the rest of the data points. We identified these extreme values using the Grubb’s test and a conservative alpha of 0.025. We did not include them in the above distribution analyses, since the analyses are very sensitive to extreme values and would make characterization of the general distribution shape difficult.

To compare the extent to which individual participants differed from the control mean among all parameters, we first calculated for each parameter the squared Z-scores for all control and MS participants relative to the control mean and standard deviation. We then averaged the squared Z-scores across parameters for each participant. Finally, we tested for differences between the control and MS distributions of average squared Z-scores using the two-tailed two-sample Kolmogorov-Smirnov test.

## 3. Results

Because the neuropathophysiology underlying clinical heterogeneity in MS is little understood and could manifest itself in different ways, we compared MS participants with controls on 16 variables that reflect components of the voluntary motor command. We examined differences in central tendency, variability, and the shape of the sample distributions.

### 3.1. Maximal strength

We first examined maximal strength of the MS group to characterize our sample. Maximal strength for each participant is displayed graphically in **Figure 3**. There were significant main effects of *Group* (F(1,95) = 9.5, *p* = 0.003) and *Torque Direction* (F(1,95) = 465.6, *p* < 0.0001) on maximal strength as well as a significant *Group x Torque Direction* (F(1,95) = 4.6, *p* = 0.034). Maximal strength was lower in the MS group compared with the control group with a mean difference of −12.2 Nm [−20.0, −4.3] on average across muscles (−6.1 Nm [−15.8, 3.5] for the TA and −18.2 Nm [−8.5, −27.8] for the soleus). For both muscles, the estimated marginal mean for maximal strength in the MS group was approximately 83% of that for the control group. The covariate of *Age_c* was also a significant predictor of maximal strength (F(1,95) = 7.8, *p* = 0.006; ß = −0.41 [−0.71, −0.12]), as was *Sex* (F(1,95) = 20.8, *p* < 0.0001; mean difference = 20.3 [11.5, 29.1]).

**Figure 3.**
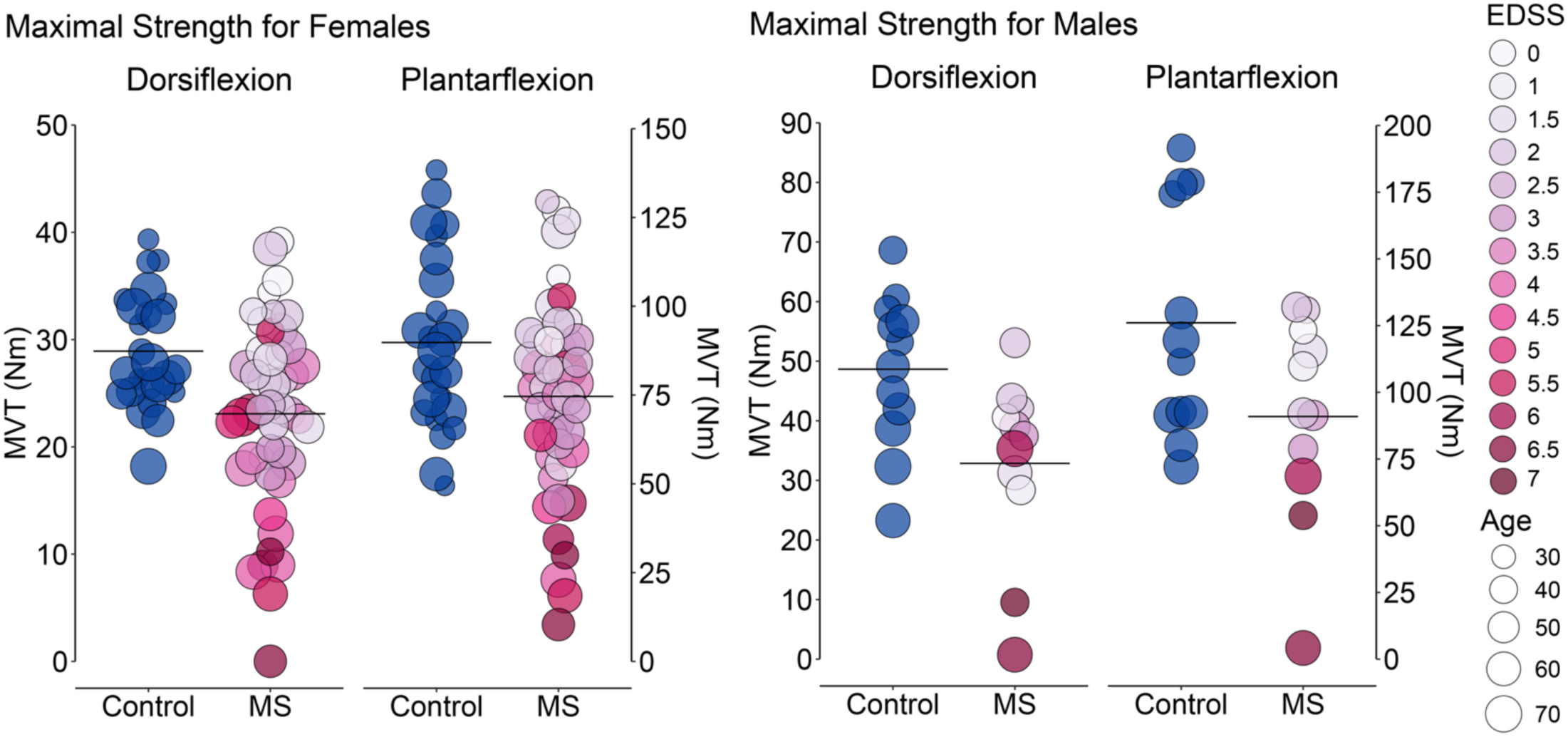
Maximal voluntary torque in dorsiflexion and plantarflexion for females (top left) and males (top right) Each circle represents an MVT value for one participant, and black lines indicate the group mean. Circle size corresponds with Age, and circle color in the MS group corresponds to the participant EDSS score.

### 3.2. Dataset characterization and individual motor unit firing profiles

In total, 14,480 decomposed motor unit spike train recordings were suitable for analysis, with an average of 16.9 per trial for the TA and 15.9 per trial for the soleus. Of these, 8,853 were designated as recordings from unique motor units, with an average number of unique motor units per participant of 37.4 for the TA and 60.0 for the soleus. The total number of spike train recordings and average number of spike trains per trial for each muscle, sex, and group combination are reported in Table 2 along with similar descriptive statistics for the number of unique motor units per participant.

**Table 2.** The mean ± SD (total) number of motor unit spike trains decomposed per trial (top) and the mean ± SD (total) number of unique motor units per participant (bottom) for each group, muscle, and sex combination.

|  | ***mean ± SD (total) number of motor unit spike trains decomposed per trial*** |  |  |  |
| --- | --- | --- | --- | --- |
|  | **MS** |  | **Control** |  |
|  | *Female (N = 47)* | *Male (N = 11)* | *Female (N = 26)* | *Male (N = 12)* |
| **TA** | 14.9* ± 7.4 (3078) | 18.8* ± 11.7 (1290) | 15.4 ± 6.9 (1615) | 23.9 ± 9.5 (1180) |
| **Soleus** | 12.8 ± 6.5 (2922) | 16.0 ± 8.4 (1065) | 16.6 ± 7.4 (2008) | 22.0 ± 12.4 (1322) |
|  | ***mean ± SD (total) number of unique motor units per participant*** |  |  |  |
|  | **MS** |  | **Control** |  |
|  | *Female (N = 47)* | *Male (N = 11)* | *Female (N = 26)* | *Male (N = 12)* |
| **TA** | 32.0* ± 18.8 (1409) | 51.1* ± 38.3 (562) | 33.5 ± 16.0 (837) | 50.6 ± 27.4 (607) |
| **SOL** | 44.0 ± 27.3 (2068) | 64.7 ± 38.4 (712) | 64.9 ± 34.7 (1623) | 86.3 ± 34.3 (1035) |

\*There were three females and one male in the MS group that did not have usable motor unit recordings for the TA. For two participants, this was due to an inability to complete the task, and for the other two, it was because of poor decomposition results. Those participants are not included in the aggregate numbers for the TA.

Participants in the MS group were generally able to perform the triangle task satisfactorily. Compared with control participants, it sometimes required more trials for MS participants to achieve sufficiently smooth torque profiles. Two participants in the MS group had little-to-no voluntary control of their dorsiflexors and were not able to complete the dorsiflexion task due to profound weakness.

There was a wide range of motor unit firing profiles in the MS group that can be appreciated visually by the examples in **Figure 4**. All three MS participants whose data is shown had both supraspinal and spinal lesions. Motor unit firing profiles from the control participant (25-29 yo) is typical for a young adult for both muscles (Beauchamp *et al*., 2022). Although S01 was of a similar age (30-34 yo) and is without motor deficits, the firing profiles in her TA motor units have noticeable differences: a compressed recruitment range, significant self-sustained firing after cessation of the task, and notable synchrony among motor unit firing patterns.

**Figure 4.**
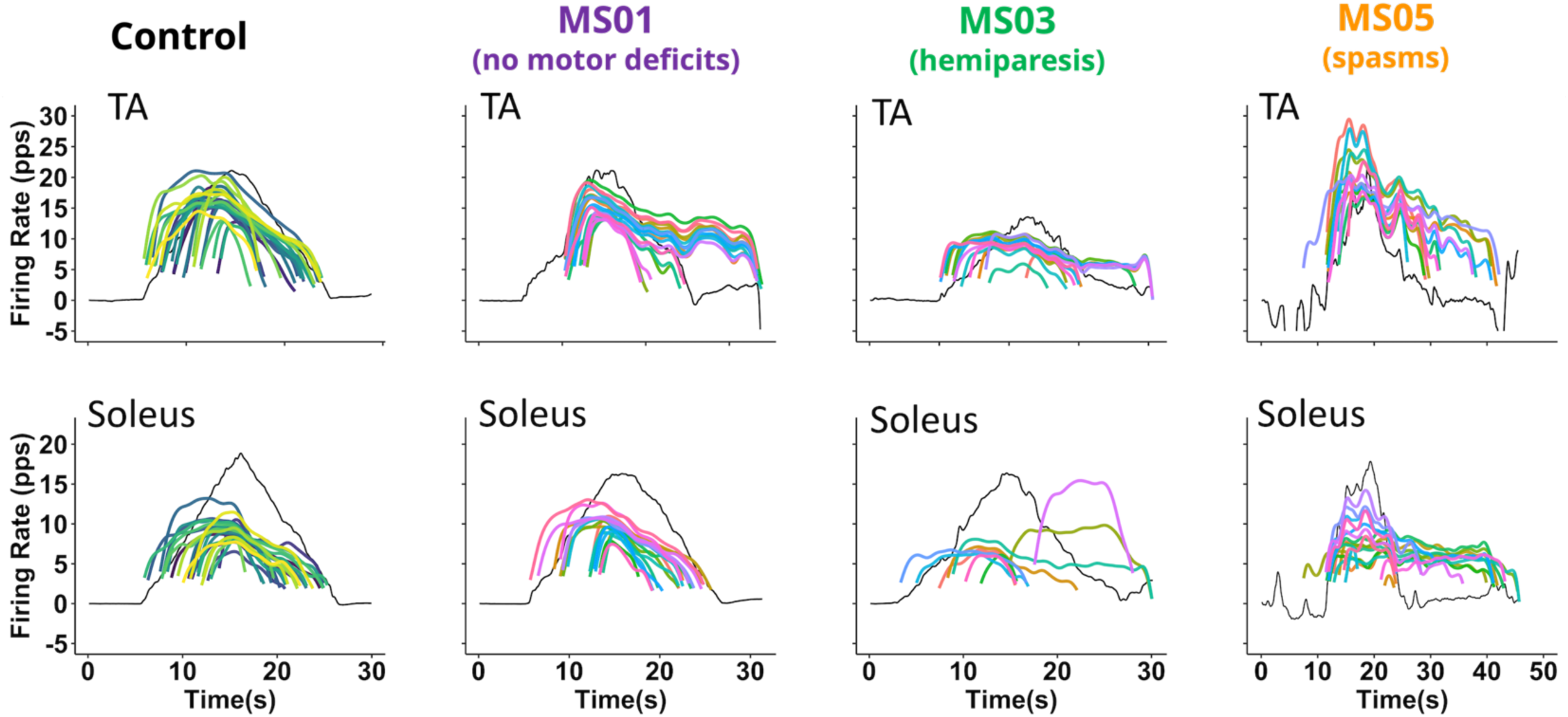
Motor unit firing patterns from single trials. Smoothed motor unit discharge rates from one control participant (left) and three MS participants are shown in color, and torque over time is shown in black. Each colored line shows the firing pattern of a different motor unit. Data from the TA during dorsiflexion are shown on the top row, and data from the soleus during plantarflexion are shown on the bottom row.

S03 was an older adult (60-64 yo) with mild hemiparesis and spasticity of the right lower extremity. Her motor units had very little change in their rates (i.e., a low rate attenuation slope) following a shallow initial acceleration, with self-sustained firing of some of the motor units after task cessation. This collection of characteristics is generally reflective of a balanced inhibition pattern and is similar to those of motor unit firing patterns of individuals with hemiparetic stroke (Mottram *et al*., 2009, 2014; Beauchamp *et al*., 2023a).

S05 was 45-49 yo, with mild bilateral weakness, left lower extremity spasticity, bilateral lower extremity spasms, and central neuropathic pain symptoms. It was difficult for her to control her torque purposefully at times, and even the anticipation of the impending triangle task caused her ankle muscles to contract unpredictably in a manner that did not occur in between trials (see baseline deviations). Her motor units had high firing rates with large acceleration amplitudes at initial recruitment and significant self-sustained firing after task cessation. This collection of characteristics is generally reflective of a loss of inhibition.

### 3.3. Differences in group means

We first tested whether the MS and control groups differed on the reverse engineering variables in terms of their group means. Given our overall hypothesis of heterogeneity in the MS group, we reasoned that either finding a difference or finding a lack thereof would be plausible. For example, there could be a range of possible delta-F differences relative to controls depending on whether neuromodulation is higher, lower, or unchanged and if the pattern of inhibition becomes more or less reciprocal or balanced. Thus, if the MS group displays heterogeneity with values that are both higher and lower than controls, then differences in group means may not emerge. Alternatively, it could be that there *are* more consistent changes in MS (either higher or lower than controls) that would be reflected in a difference in group means.

Participant means for the reverse engineering variables are shown in **Figure 5** broken down by Group and Muscle. A summary of the linear mixed models results for fixed effects of interest (*Group, Muscle, Group x Muscle*) for each reverse engineering variable is shown in **Table 3**. A similar summary for the covariates (*Age_c, Sex, TQ_recrt_c*) is shown in **Table S1**.

**Figure 5.**
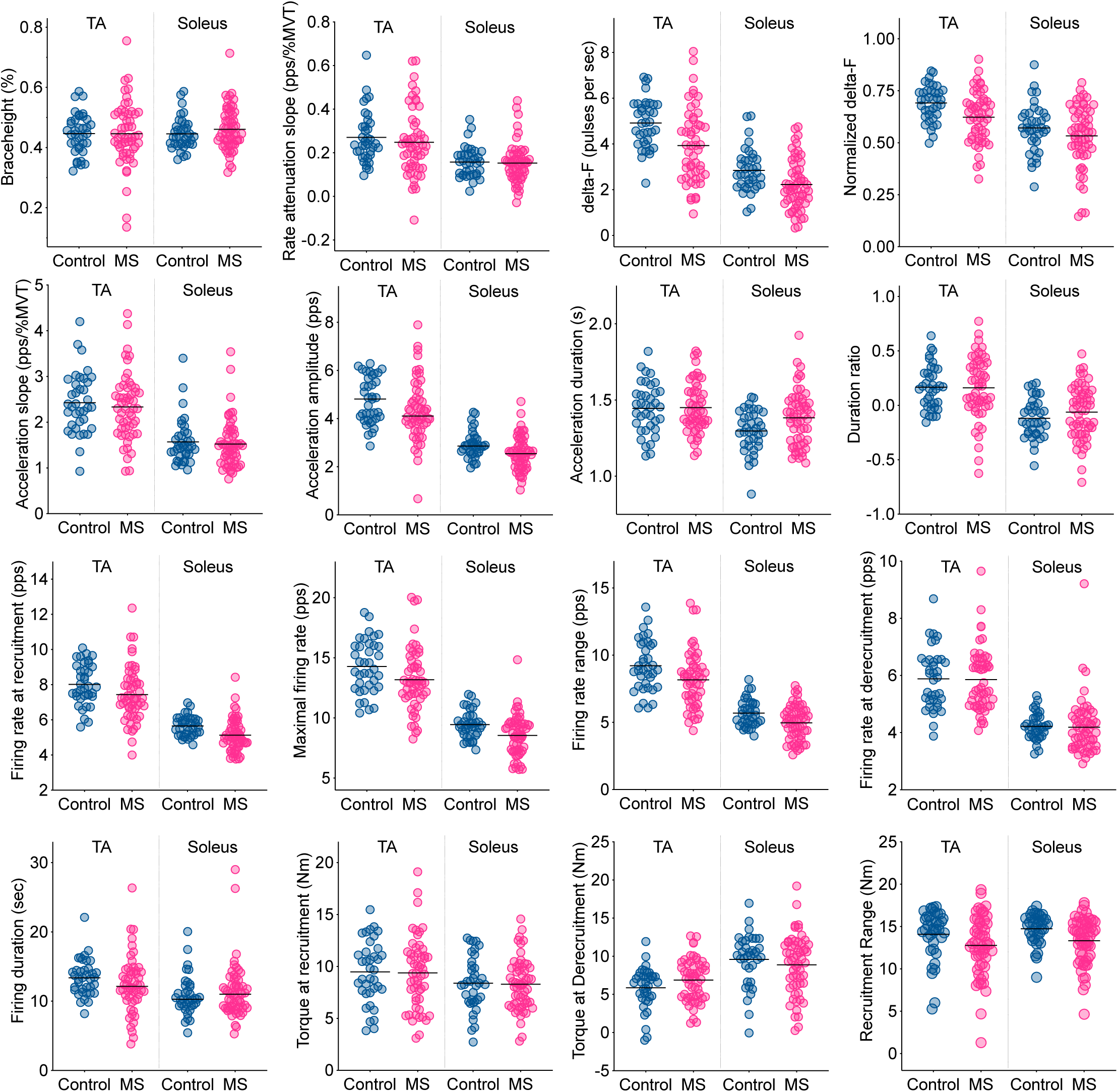
Participant values for reverse engineering variables. Median values for control participants are shown as blue circles, and those for MS participants are shown as pink circles. Group mean values for each *Group* and *Muscle* combination are indicated by horizontal black lines. Group mean values were significantly lower for the MS group for delta-F, normalized delta-F, acceleration amplitude, firing rate at recruitment, firing rate range, maximal firing rate, and recruitment range (**Table 3**).

**Table 3.** Linear mixed effect model results for each reverse engineering variable: *Group, Muscle, Group x Muscle*.

|  | Fixed Effects |  |  |  |  |  |  |  |  |
| --- | --- | --- | --- | --- | --- | --- | --- | --- | --- |
|  | *Group* |  |  | *Muscle* |  |  | *Group x Muscle* |  |  |
|  | Bartlett | *p* | mean diff<br>[CI] | Bartlett | *p* | mean diff<br>[CI] | Bartlett | *p* | TA: mean diff<br>[CI]<br>SOL: mean<br>diff<br>[CI] |
| Delta-F | 9.7 | **0.002** | -0.62<br>[-1.01, -0.24] | 115.1 | **< 0.0001** | 1.90<br>[1.66, 2.14] | 1.6 | 0.21 | -0.31<br>[-0.78, 0.17] |
| Norm<br>delta-F | 6.8 | **0.009** | -0.05<br>[-0.08, -0.01] | 17.5 | **< 0.0001** | 0.12<br>[0.09, 0.15] | 0.82 | 0.36 | -0.02<br>[-0.07, 0.02] |
| Duration<br>Ratio | 1.0 | 0.31 | 0.04<br>[-0.03, 0.10] | 64.6 | **< 0.0001** | 0.24<br>[0.19, 0.29] | 1.2 | 0.28 | -0.06<br>[-0.16, 0.05] |
| Braceheight | 0.78 | 0.38 | 0.01<br>[-0.01, 0.03] | 3.1 | 0.080 | -0.02<br>[-0.03, 0.002] | 0.85 | 0.36 | -0.02<br>[-0.05, 0.02] |
| Acceleration<br>amplitude | 4.9 | **0.028** | -0.32<br>[-0.59, -0.04] | 118.0 | **< 0.0001** | 1.81<br>[1.59, 2.02] | 0.19 | 0.66 | -0.10<br>[-0.53, 0.34] |
| Acceleration<br>slope | 0.03 | 0.85 | -0.02<br>[-0.21, 0.18] | 72.5 | **< 0.0001** | 0.85<br>[0.69, 1.00] | 0.49 | 0.48 | -0.11<br>[-0.42, 0.20] |
| Acceleration<br>duration | 0.22 | 0.64 | 0.01<br>[-0.03, 0.06] | 20.7 | **< 0.0001** | 0.09<br>[0.05, 0.13] | 0.46 | 0.50 | -0.03<br>[-0.10, 0.05] |
| Rate<br>attenuation<br>slope | 0.33 | 0.57 | -0.01<br>[-0.04, 0.02] | 38.6 | **< 0.0001** | 0.09<br>[0.06, 0.12] | 0.09 | 0.77 | -0.01<br>[-0.06, 0.04] |
| Firing rate at<br>recruitment | 5.8 | **0.016** | -0.49<br>[-0.89, -0.10] | 127.6 | **< 0.0001** | 2.25<br>[1.99, 2.50] | 0.08 | 0.77 | -0.08<br>[-0.58, 0.43] |
| Firing rate<br>range | 7.5 | **0.006** | -0.75<br>[-1.26, -0.24] | 134.2 | **< 0.0001** | 3.42<br>[3.03, 3.80] | 1.14 | 0.29 | -0.44<br>[-1.20, 0.33] |
| Maximal<br>firing rate | 6.1 | **0.013** | -0.96<br>[-1.63, -0.28] | 58.7 | **< 0.0001** | 4.76<br>[4.30, 5.23] | 0.56 | 0.46 | -0.36<br>[-1.29, 0.58] |
| Firing rate at<br>de-recruitment | 0.41 | 0.52 | -0.10<br>[-0.40, 0.20] | 109.2 | **< 0.0001** | 1.54<br>[1.33, 1.74] | 0.06 | 0.80 | 0.05<br>[-0.36, 0.47] |
| Recruitment<br>torque | 0.18 | 0.68 | -0.15<br>[-0.85, 0.54] | 9.1 | **0.003** | 0.97<br>[0.36, 1.58] | <0.0001 | 0.99 | 0.002<br>[-1.22, 1.21] |
| De-recruitment<br>torque | 0.11 | 0.75 | 0.16<br>[-0.76, 1.07] | 33.3 | **< 0.0001** | -2.89<br>[-3.56, -2.22] | 4.2 | **0.041** | 1.35<br>[0.10, 2.61] |
| Duration | 0.37 | 0.54 | 0.39<br>[-0.84, 1.64] | 10.6 | **0.001** | 2.56<br>[1.37, 3.78] | 1.2 | 0.54 | -0.49<br>[-1.98, 0.99] |
| Recruitment<br>Range | 6.6 | **0.010** | -1.21<br>[-2.10, -0.33] | 0.27 | 0.60 | -0.28<br>[-1.42, 0.88] | 0.34 | 0.56 | 0.42<br>[-1.00, 1.84] |

There was a significant effect of *Group* for six of the 16 variables. Delta-F, normalized delta-F, acceleration amplitude, firing rate at recruitment, firing rate range, maximal firing rate, and recruitment range were all lower in the MS group compared with the Control group (mean differences shown in **Table 3**). In other words, on average, motor units from participants in the MS group were recruited at a lower firing rate, their initial acceleration was smaller in amplitude, they reached a lower maximal firing rate and exhibited less overall modulation of the firing rate, and they experienced less recruitment hysteresis. These findings indicate that most of the participants with MS demonstrated motor unit firing patterns similar to those from S03 shown in **Figure 4**.

There was a significant effect of *Muscle* for all parameters except for recruitment range and braceheight. Delta-F, normalized delta-F, duration ratio, acceleration amplitude, acceleration slope, acceleration duration, rate attenuation slope, initial firing rate, maximal firing rate, firing rate range, final firing rate, duration, and torque at recruitment were all higher in the TA than in the soleus, on average across groups, whereas torque at de-recruitment was lower in the TA than in the soleus. The *Group* x *Muscle* interaction was significant for only torque at de-recruitment. The between-muscle difference in torque at de-recruitment was smaller for the MS group than for the control group (estimated difference: 2.22 [1.39, 3.05] vs. 3.57 [2.59, 4.55]).

There was a significant effect of *TQ_recrt_c* for all variables except delta-F (ß = 0.005 [−0.004, 0.014], *p* = 0.30). Of these, duration ratio, acceleration slope, firing rate at recruitment, firing rate at de-recruitment, torque at de-recruitment, and recruitment range had a positive relationship with *TQ_recrt_c*. Normalized delta-F, acceleration amplitude, acceleration duration, firing rate range, maximal firing rate, and duration had a negative relationship with *TQ_recrt_c*.

*Age_c* had a significant effect on several of the variables. With increasing Age, there were decreases in delta-F, normalized delta-F, duration ratio, acceleration amplitude, firing rate range, firing rate at recruitment, firing rate range, maximal firing rate, and torque at recruitment, whereas there was an increase in firing rate at de-recruitment. These findings are consistent with previous studies examining the effect of age on some of these parameters in elbow muscles (Hassan *et al*., 2021), ankle muscles (Orssatto *et al*., 2021), and knee muscles (Guo *et al*., 2024). The effect of *Sex* was significant only for acceleration amplitude (ß = 4.6, *p* = 0.033, mean difference: −0.31 [−0.58, −0.03] and acceleration slope (ß = 4.3, *p* = 0.038, mean difference: −0.24 [−0.45, −0.03]).

### 3.4. Group differences in sample distribution shape

To compare how the MS and control samples were distributed around their means, we examined characteristics of the shapes of the distributions and the extent to which individual participants differed from the control mean.

When testing for group-related statistical differences in distribution shape (two-sample Komolgorov-Smirnov test on the group mean-centered distributions) we found no significant differences for any of the parameters for either of the muscles, perhaps due to a lack of power or sensitivity for this particular test (D ranging from 0.08 to 0.26 and *p*-values ranging from 0.99 to 0.09).

Nevertheless, we found intriguing differences between the MS and control distribution shapes in terms of variability and dispersion that varied among parameters. These are shown in **Figure 6A** as the ratios of the interquartile range and range for each parameter (MS group relative to controls) along with the coefficients of variation (**Figure 6B**) and group distributions of the average squared Z-score across parameters (**Figure 6C**). We detail findings in this section that indicate that (1) qualitatively, the two groups are distributed differently on most of the reverse engineering parameters, (2) not everyone in the MS group differs from the control group in the same way, and (3) on average across parameters, participants in the MS group deviated from the control group mean more than those in the control group.

**Figure 6.**
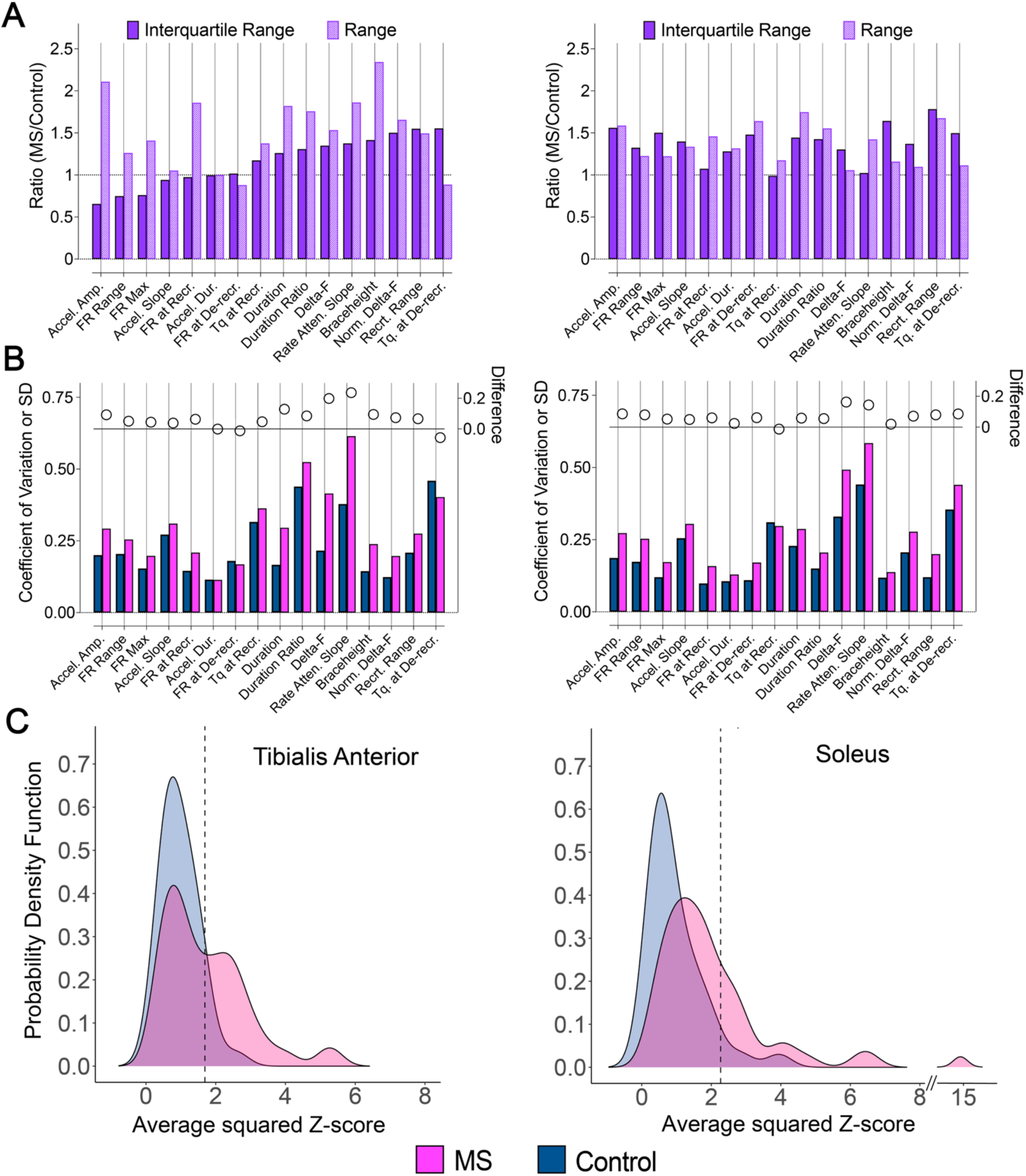
Distributional characteristics for the reverse engineering parameters for MS and control groups for the TA (left column) and soleus (right column) **A.** Between-group ratios (MS/Control) of the interquartile range and the overall range. **B.** The coefficients of variation for each parameter for MS (pink) and control (blue) groups (bottom) and the difference in coefficients of variation between groups (top). The values shown for duration ratio are standard deviations. **C.** Probability density functions of the average squared Z-score (across parameters per participant), for all participants.

#### 3.4.1. Comparison of sample distribution shapes for the tibialis anterior

For the TA, the MS group had a wider range of values than the control group (by 1.05 – 2.34x) for all parameters except firing rate at de-recruitment, torque at de-recruitment, and acceleration duration. The relative interquartile range for the MS group compared with controls varied among parameters.

For three of the parameters (maximal firing rate, firing rate range, acceleration amplitude), the MS distributions had a narrower center peak than the control distribution (i.e., a smaller interquartile range) in combination with a larger overall range. This pattern indicated that (1) most MS participants were clustered at the center of the distribution and had parameter values that were more similar to each other than control participants at the center of the control group distribution, and (2) that the other MS participants had more extreme values than the control group. Firing rate at recruitment and acceleration slope for the MS group had a similar interquartile range to that of controls, whereas the interquartile range for the MS group was greater for the remaining parameters.

The coefficients of variation (and variance for duration ratio) were higher for the MS group for all parameters except acceleration duration, firing rate at de-recruitment, and torque at de-recruitment. Of the parameters with higher coefficients of variation, the variance of the MS group was statistically higher for delta-F (2.7 vs. 1.1; F(1,88) = 7.4, *p* = 0.007), normalized delta-F (0.015 vs. 0.007; F(1,88) = 5.2, *p* = 0.033), duration ratio (0.083 vs. 0.040; F(1,89) = 4.3, *p* = 0.04), and braceheight (0.011 vs. 0.004; F(1,89) = 4.1, *p* = 0.046). The variances for the MS group were approximately 2 to 3 times those of controls.

One possible reason for the increased variability and wider ranges in the MS group is simply that we included participants with a range of impairment levels. However, this pattern occurred in the context of MS distributions that had substantial dispersion both higher *and* lower than those of controls. This occurred for variables with a group mean difference (e.g., delta-F), as well as those without a group mean difference (e.g., braceheight). If our sample of participants with MS had the same underlying pathophysiology with differing severity, we would expect MS participant values to be shifted either higher or lower than the control mean but not both. Furthermore, the MS distributions for some parameters appeared to have multiple peaks, with a collection of participant values centered higher than the control mean, and a collection of participant values centered lower than the control mean. This pattern was most pronounced for braceheight, duration ratio, firing rate at de-recruitment, torque at recruitment, torque at de-recruitment, delta-F, and normalized delta-F. These apparent and/or emerging bimodal distributions could be interpreted as evidence of different physiological phenotypes.

#### 3.4.2. Comparison of sample distribution shapes for the soleus

For the MS distributions for the soleus, the most common pattern of dispersion among parameters was that of a larger or comparable interquartile range and a larger overall range.

The coefficient of variation (and variance for duration ratio) was higher for the MS group for all parameters except for torque at recruitment. Of those parameters, the variance of the MS group was statistically higher than that of the control group for duration ratio (0.06 vs. 0.03; F(1,93) = 4.4, *p* = 0.04), maximal firing rate (2.8 vs. 1.3; F(1,93) = 4.5, *p* = 0.036), firing rate at recruitment (0.83 vs. 0.31; F(1,93) = 4.6, *p* = 0.034), firing rate at de-recruitment (0.93 vs. 0.21; F(1,93) = 4.7, *p* = 0.032), and recruitment range (7.0 vs. 3.2; F(1,93) = 5.0, *p* = 0.028). The variances for the MS group were approximately 2 to 4 times those of controls.

#### 3.4.3. Overall magnitude of variation from control means among parameters

We next summarized the extent to which participants in the MS group deviated (both higher and lower) from control group means for all parameters, even those without group mean differences. **Figure 5C** shows the group distributions for each participant’s average squared Z-score, displayed for each group and muscle.

The distributions were significantly different between groups for both muscles (TA: D = 0.41, *p* = 0.0009; soleus: D = 0.44, *p* = 0.0002), with median values of 0.88 (TA) and 0.69 (soleus) for the control group and 1.50 (TA) and 1.64 (soleus) for the MS group. For the TA, 46.3% of the MS participants had an average squared Z-score value that was greater than the 95% percentile of the control distribution. This was the case for 29.3% of the MS participants for the soleus. This finding indicates that MS participants tended to deviate from the control mean (either higher or lower) more than control participants, even when including variables for which there were no group mean differences.

## 4. Discussion

We identified pathological aspects of excitatory, inhibitory, and neuromodulatory components of voluntary motor commands at the level of the spinal motoneuron in a clinically heterogeneous sample of patients with MS. We leveraged recordings of discharge patterns of many single motor units by applying a reverse engineering paradigm to characterize the synaptic inputs that generated those discharge patterns in the spinal cord. This approach allowed us to examine neural control of voluntary movement in people with MS with more depth than previously possible.

Our overall scientific hypothesis was that multiple neurophysiological phenotypes of voluntary motor command pathology are evident in the MS population. Our rationale derives from the different pathologies of the voluntary motor command found to date in people with spinal cord injury and people with hemiparetic stroke. People with MS unpredictably develop motor deficits typical of one, both, or neither of these populations, and they can have damage to the brain and/or spinal cord. Therefore, it is plausible that voluntary motor command pathology manifests differently in different patients.

There are two main findings from this study. First, we generally observed that MS values for many of our reverse engineering parameters were highly variable. The parameter values of some participants were abnormally high relative to the control mean, whereas others were abnormally low.

Second, for seven parameters, the MS group mean values were statistically significantly lower than those of controls. However, the MS distributions for these parameters also displayed the variability discussed above, often including extreme MS values that were *higher* than any control values. Aside from recruitment range, these parameters (maximal firing rate, firing rate range, firing rate at recruitment, acceleration amplitude, delta-F, normalized delta-F) are associated with motoneuron excitability and could be impacted by either the level of neuromodulation or the pattern of inhibition (Powers & Heckman, 2017). Thus, decreased motoneuron excitability *during voluntary contractions* was the most common, but not the only, finding in the MS group. Taken together, these findings support our overall hypothesis of multiple neurophysiological phenotypes within the MS population and warrant further exploration in future research.

Decreased motoneuron excitability in MS seems counterintuitive given that people with CNS injury involving upper motoneurons typically present with *increased* spinal excitability at rest (i.e., when not moving). Indeed, that was the case for many of our participants with MS, who had motor unit firing patterns consistent with decreased motoneuron excitability during voluntary contractions but also presented, at rest, with increased deep tendon reflexes and/or the presence of multi-joint spasms, spasticity, substantial muscle twitches in response to sensory input, and/or cramps. This combination is possible given that there are different neural inputs to the motoneuron pool (and thus different mixtures of neuromodulation and inhibition) during voluntary contractions compared with the resting state, and there is evidence that this pattern also appears in patients with hemiparetic stroke (Mottram *et al*., 2009, 2014; McPherson *et al*., 2018d, 2018c; Beauchamp *et al*., 2023a).

Previous research investigating mechanisms underlying motor deficits in MS have focused almost exclusively on the excitatory component of the voluntary motor command. They have assessed intracortical, corticospinal, and resting spinal reflex excitability, typically using transcranial magnetic stimulation and evoked H-reflexes, often within the context of spasticity rather than deficits in voluntary movement (e.g., Houdayer *et al*., 2015). A more comprehensive picture of the voluntary motor command has emerged for the hemiparetic stroke and spinal cord injury populations, as a small yet growing collection of studies have broadened the focus beyond excitation to investigate dysfunctional neuromodulatory and inhibitory inputs to motoneurons (Gorassini *et al*., 2004; Norton *et al*., 2008; Mottram *et al*., 2009, 2014; Murray *et al*., 2010, 2011b; D’Amico *et al*., 2013; McPherson *et al*., 2018c, 2018b; Beauchamp *et al*., 2023b, 2023a, 2024b; Mahrous *et al*., 2024), although full characterizations of reverse engineering parameters in these populations are in progress. In subsequent sections, we will briefly discuss the current body of knowledge in spinal cord injury and hemiparetic stroke with respect to its relevance to the MS population and our current findings.

### 4.1. Motor deficits in spinal cord injury: loss of descending excitation and neuromodulation resulting in weakness and unregulated sensory transmission

Common sensorimotor deficits in incomplete spinal cord injury can emerge in patients with MS (Joy *et al*., 2001), appearing unilaterally or bilaterally and affecting the upper and/or lower extremity. They include weakness, spasticity, loss of somatosensation, neuropathic pain, and strong, involuntary, multi-joint spasms that are evoked by changes in somatosensory input, vestibular input, and/or sleep/wake arousal (Mahrous *et al*., 2023; Moreno Romero *et al*., 2024). All three components of the motor command are affected in spinal cord injury, collectively resulting in reduced voluntary motor output and unregulated sensory afferent transmission (Mahrous *et al*., 2023).

Losses of both excitatory and neuromodulatory input to motoneurons after spinal cord injury limit the ability to voluntarily activate motoneurons. Excitatory input, which initiates and controls voluntary movement, is diminished because of damage to corticospinal, reticulospinal, and vestibulospinal tracts (Baker, 2011). Neuromodulatory input, which increases motoneuron excitability to effectively transform the excitatory input into sustained motoneuron discharge, is reduced because of damage to raphespinal and ceruleospinal tracts (Mahrous *et al*., 2023). Acutely, the loss of neuromodulatory input to motoneurons results in profoundly unexcitable motoneurons and the associated clinical manifestations of spinal shock, areflexia, and paralysis (Heckman *et al*., 2009; Murray *et al*., 2010). In the months following the injury, motoneuron excitability recovers paradoxically despite the continued absence of serotonin and norepinephrine. Such recovery results from upregulation of constitutively active serotonin (5-HT_2c_) and norepinephrine receptors (⍺_1_) on motoneuron dendrites that facilitate PICs in the absence of the neurotransmitters (Murray *et al*., 2010, 2011a; Rank *et al*., 2011; D’Amico *et al*., 2013).

While restoration of PICs via adaptation of monoaminergic receptors is vital for recovery of some voluntary muscle activation in spinal cord injury (Murray *et al*., 2010), weakness persists due to the continued loss of descending excitatory input. In addition, a notable side effect of PIC restoration is the emergence of profound muscle spasms and hyperreflexia. Inhibitory input to motoneurons and the capacity for post-synaptic inhibitory currents on motoneuron dendrites are both drastically decreased and thus cannot appropriately offset the restored motoneuron excitability (Boulenguez *et al*., 2010; Caron *et al*., 2020; Mahrous *et al*., 2024). There is also a substantial increase in non-purposeful excitatory input to motoneurons from unregulated sensory afferent transmission. In the intact nervous system, there is tight descending modulation of spinal afferent transmission to allow only salient information to reach the motoneuron (Eccles & Lundberg, 1959; Lemon, 2008). Damage to descending pathways interferes with their activation of inhibitory interneurons that typically gate sensory transmission to motoneurons in a task-appropriate manner (Seki & Fetz, 2012; Kalambogias & Yoshida, 2021).

### 4.2. Is there evidence of damage to descending neuromodulatory pathways in MS?

It is plausible that reductions in descending serotonin and norepinephrine and the adaptive processes that follow in spinal cord injury could also underlie weakness, spasms, and hyperreflexia in some patients with MS.

The structure and function of brainstem-spinal monoaminergic pathways in MS was investigated systematically by White and others in a series of studies during the 1970s, 80s, and 90s using rats and guinea pigs with experimental autoimmune encephalomyelitis (EAE), the animal model of MS. Collectively, these studies provided evidence of morphological damage to descending monoaminergic axons and axon terminals (Bieger & White, 1981; White *et al*., 1985, 1989, 1990; White & Bowker, 1988; Vyas *et al*., 1988), reduced levels of serotonin and norepinephrine in the spinal gray matter (Lycke & Roos, 1973; White *et al*., 1983, 1990; Krenger *et al*., 1986; Samathanam *et al*., 1991), and reduced serotonin receptor sensitivity in the spinal cord (White, 1979; White & Bieger, 1980) of animals with EAE. There is evidence that such changes are specific to monoaminergic pathways, presumably because their small diameter and thin or lack of myelination make them more susceptible to injury (White & Bowker, 1988; Polak *et al*., 2011), and thus affect not only serotonin and norepinephrine but the neurotransmitters that are co-released (e.g., substance-P, enkephalins, BDNF, and other peptides) (Hökfelt *et al*., 2000; Hodges & Richerson, 2008; Caramia *et al*., 2023).

Morphological changes to the descending monoaminergic pathways visualized using histofluorescence and immunohistochemistry included gross distortions in shape with periodic areas of “balloon-like” swelling where the axons encountered blood vessels with perivascular inflammatory cuffs (Bieger & White, 1981; White *et al*., 1985; White & Bowker, 1988). There was evidence of a build-up of serotonin and norepinephrine in the areas of axonal swelling and a loss of noradrenergic and serotonergic terminals in the ventral horn, suggesting that the swelling impaired the axoplasmic flow required to transport the neurotransmitters to their synaptic terminals (Bieger & White, 1981; White *et al*., 1985). Decreases in norepinephrine and dopamine beta-hydroxylase, the enzyme crucial for norepinephrine synthesis, were also found in the spinal cord of mice with EAE in two more recent studies, along with evidence of glial inflammation (i.e., increases in glial fibrillary acidic protein) (Polak *et al*., 2011; Torrillas-de La Cal *et al*., 2023).

Importantly, this body of work demonstrated that the observed changes to the descending monoaminergic system were often associated with clinical symptoms. First, a rostrocaudal gradient in changes was consistently found, such that the lumbosacral spinal cord was the most affected (White *et al*., 1983, 1985, 1989; Krenger *et al*., 1986, 1989). The authors attributed this pattern to the increasing probability of encountering collections of inflammatory cells as axons of descending monoaminergic pathways traverse farther through the spinal cord. This finding is consistent with the pattern of sensorimotor symptoms of EAE animals as well as patients with MS, as the lower extremities are typically more affected (Joy *et al*., 2001).

Second, axonal deformities in descending monoaminergic neurons in EAE animals appeared with the earliest motor symptoms and were correlated with both the level of perivascular inflammation and the severity of hindlimb paralysis (White *et al*., 1990). These findings are consistent with reductions in serotonin turnover found in the CSF of patients with MS, which were more pronounced with patients with more disability (Andersen *et al*., 1981; Markianos *et al*., 2009). In the cervical and thoracic spinal cord, axon terminal morphological changes (White *et al*., 1985) and depletion of serotonin and norepinephrine (Krenger *et al*., 1989) improved following recovery from paralysis, with the authors suggesting that apparent reinnervation of affected areas could result from sprouting of undamaged axons and axon terminals. However, no improvements were found in the lumbosacral region of the spinal cord, as the initial disturbances persisted and sometimes worsened with subsequent relapses (White *et al*., 1989, 1990; Krenger *et al*., 1989). In contrast, levels of spinal glutamate and GABA that were reduced during an attack fully normalized after recovery at all levels of the spinal cord (Honegger *et al*., 1989).

In further support of alterations to the monoaminergic system in MS, *ascending* noradrenergic, serotonergic, dopaminergic projections from the brainstem to the brain have also been of interest in the MS field because of the postulated contribution of their disruption to prevalent clinical symptoms of fatigue, depression, and cognitive dysfunction (Feinstein *et al*., 2016; Manjaly *et al*., 2019; Carandini *et al*., 2021a). Collectively, findings from studies using mice with EAE and patients with MS (Gadea *et al*., 2004; Polak *et al*., 2011; Carandini *et al*., 2021c, 2021b) indicate that ascending monoaminergic transmission is less effective due to both structural damage to ascending monoaminergic pathways and an inflammation-induced reduction in monoamine synthesis (Manjaly *et al*., 2019; Carandini *et al*., 2021a). Some studies have shown relationships between various measures of ascending monoaminergic pathway structure and/or function with clinical symptoms of fatigue (Carandini *et al*., 2021c), depressive symptoms (Carotenuto *et al*., 2020), and cognitive dysfunction (Gadea *et al*., 2004; Carotenuto *et al*., 2020). Inflammatory lesions found in the raphe nuclei of patients with MS via MRI are also consistent with changes in monoaminergic signaling in MS (Nguyen *et al*., 2021).

If there are substantial reductions in serotonergic and noradrenergic input to motoneurons in MS, they would assuredly contribute to weakness, at least initially. It is an open question whether compensatory upregulation of constitutively active serotonin and norepinephrine receptors on motoneuron dendrites occurs as in spinal cord injury. In support of this idea, 36% of our participants with MS reported spinal cord injury-like spasms, which are not typically found in other neurological populations. However, some of these participants had motor unit firing patterns with clear non-linearities consistent with PICs and others had extremely linear motor unit firing patterns. These observations indicate that perhaps some patients with MS experience adaptation of motoneuronal serotonin receptors and others do not.

### 4.3. Motor deficits in stroke: loss of corticospinal excitation, increased reticulospinal excitation and neuromodulation, and pathological inhibition

Common sensorimotor deficits in hemiparetic stroke can also emerge in patients with MS (Joy *et al*., 2001). In hemiparetic stroke, they result largely from injury to the densely-packed descending corticospinal and ascending somatosensory pathways traveling through the internal capsule. They include the following impairments, presenting unilaterally in both the upper and lower extremity: weakness, loss of fine motor control, spasticity and increased resting muscle tone, diminished somatosensation, and abnormal flexor and extensor synergies that limit the ability to control proximal and distal joints independently (Li *et al*., 2019; McPherson & Dewald, 2019, 2022).

In the intact nervous system, lateral corticospinal and reticulospinal systems work in parallel to deliver excitatory input to motoneurons to produce fine, fractionated movements (corticospinal) with appropriate postural stability (reticulospinal) (Baker, 2011; Baker *et al*., 2015). In addition to its excitatory fibers, the reticulospinal system also includes the raphespinal and ceruleospinal tracts, and therefore also delivers neuromodulatory input to motoneurons (Veasey *et al*., 1995; Schwarz *et al*., 2008; Heckman *et al*., 2009; McPherson *et al*., 2018b).

Supraspinal injury to the corticospinal tract has both direct and indirect effects on synaptic inputs to motoneurons. The loss of corticospinal excitatory input results directly in weakness and a loss of fine motor control that is supported uniquely by the sophistication of the corticospinal tract. Indirectly, there is an increased influence of diffusely-projecting excitatory input due to compensatory upregulation of the reticulospinal system, recruited via the contralesional hemisphere when task demands exhaust the ipsilesional corticospinal resources that remain (McPherson *et al*., 2018a; Karbasforoushan *et al*., 2019; McPherson & Dewald, 2022; Mooney *et al*., 2025). Reticulospinal excitatory input enables or assists with voluntary muscle activation but only within stereotyped whole-limb flexion and extension patterns (abnormal synergies) due to the pathway’s anatomical constraints (McPherson *et al*., 2018a; McPherson & Dewald, 2019, 2022). In concert, reticulospinal *neuromodulatory* input to motoneurons is also upregulated in hemiparetic stroke, contributing to increased resting muscle tone, hyperreflexia and spasticity, and abnormal synergies, through facilitation of *excitatory* reticulospinal input (McPherson *et al*., 2018c, 2018b; Beauchamp *et al*., 2024b).

There is evidence suggesting that pathological inhibitory input to motoneurons also plays a role in post-stroke weakness, despite the presence of increased spinal excitability at rest. Specifically, limited or negative motor unit rate modulation during an increasing contraction is a prominent characteristic of motor unit firing patterns in people with hemiparetic stroke (Gemperline *et al*., 1995; Chou *et al*., 2013; Mottram *et al*., 2014; Miller *et al*., 2014; Beauchamp *et al*., 2023a). Such diminished rate modulation cannot be explained by loss of excitatory input, because recruitment of additional motor units continues. Based on the reverse engineering paradigm we use (Powers *et al*., 2012; Beauchamp *et al*., 2023b; Chardon *et al*., 2023), there is increasing evidence that such a phenomenon is produced by a proportional or “balanced” pattern of inhibition (that scales proportionally to excitation), thus “blunting” the effectiveness of the excitation and contributing to weakness.

### 4.4. Is there evidence of an upregulated reticulospinal pathway in MS?

Many of our participants with MS had motor deficits consistent with an increased influence of the reticulospinal tract, as is seen after hemiparetic stroke. Most commonly, we saw a unilateral lower extremity extension synergy pattern during ambulation. This pattern was sometimes accompanied by flexion posturing in the ipsilateral upper extremity, especially for those who were the most impaired overall. Similarly, for more impaired participants, we observed associated reactions, whereby a participant’s upper extremity would move in a flexion synergy pattern during maximal efforts at the ankle.

In general, the abnormal synergy patterns seen in our participants with MS were relatively minor compared with the hemiparetic stroke population. This difference is perhaps not surprising, given that white matter lesions in MS are typically smaller and more disseminated than those with hemiparetic stroke. Therefore, focal loss of corticospinal fibers within subcortical white matter (that contributes strongly to post-stroke motor deficits) may be less extensive in MS, especially in the modern era of disease-modifying therapies that are highly effective in limiting the formation of new lesions.

### 4.5. Potential clinical interventions tailored to pathology of the voluntary motor command

There is a growing number of emerging interventions in the neurorehabilitation field that use neurostimulation to augment the effects of physical rehabilitation in people with neurological injury (Dietz & Fouad, 2014; Thompson & Wolpaw, 2015; Van Den Brand *et al*., 2015; Micera *et al*., 2020; Vose *et al*., 2022; Asan *et al*., 2022; Anderson *et al*., 2022; Wolpaw & Thompson, 2023; Moreno Romero *et al*., 2024; Kilic *et al*., 2025). Very few of these interventions have been meaningfully pursued in patients with MS, and there are two reasons why their direct transfer to the MS population may be less successful: (1) they predominantly focus on restoration of excitatory synaptic input to motoneurons without considering how altered neuromodulatory and/or inhibitory input affects motor deficits, and (2) their development is not personalized to each patient’s pathophysiology, which may be a unique need for the MS population.

Currently, the primary aim of neurostimulation interventions is typically to increase excitatory drive to motoneuron pools. They do this by facilitating transmission through damaged descending excitatory pathways and/or activating spinal circuits that support complex motor behaviors (e.g., locomotion). The most common approach is to use electrical and/or magnetic stimulation of the brain, spinal cord, and/or sensory afferents using principles of activity-dependent plasticity. Other interventions, like acute intermittent hypoxia, elicit plasticity of excitatory pathways in other ways.

With our growing understanding of the role and importance of inhibitory and neuromodulatory inputs to motoneurons in typical and pathological motor control, interventions that target these components of the motor command may be necessary to optimize outcomes. For example, an approach that diminishes unwanted proportional inhibition may be more effective in restoring motor output than those aiming to increase excitation alone. Similarly, restoring sufficient inhibitory inputs to motoneurons could prevent the occurrence of debilitating multi-joint muscle spasms that interfere with both voluntary movement and quality of life.

The utility of increasing descending neuromodulation has been demonstrated in two studies of mice with EAE. One study used prolonged daily intermittent stimulation of the nucleus raphe magnus using a wireless electrical stimulator to increase spinal serotonin (Hentall *et al*., 2006; Madsen *et al*., 2017). Stimulation was started after the initial peak of symptom worsening and resulted in improved clinical motor signs (from complete to partial hindlimb paralysis, on average) as well as a reduced number of infiltrating immune cells (by 50%), an increased number of myelinated axons (by two times), and a more favorable gene expression profile, all in the spinal cord, compared with animals with sham stimulation.

Another study used chemogenetic activation of noradrenergic neurons in the locus ceruleous (Torrillas-de La Cal *et al*., 2023), which had similar benefits as nucleus raphe magnus stimulation. Compared with animals without intervention, locus ceruleous activation from the onset of symptoms resulted in markedly less severe clinical motor deficits as well as fewer infiltrating immune cells, perivascular cuffs, demyelination, and glial activation, and a smaller loss of the key enzyme that produces norepinephrine, all in the spinal cord. The effects of locus ceruleous activation were more modest when initiated at the peak rather than the onset of symptoms.

Of course, the improvements in EAE clinical signs and disease pathology resulting from increased serotonin and norepinephrine can likely be attributed to the widespread anti-inflammatory and neuroprotective effects of these neurotransmitters; however, their restoration of neuromodulatory action on motoneurons undoubtedly contributes.

Our findings in this study are a long way off from guiding treatment selection, but if there are multiple neurophysiological “phenotypes” in MS, that would suggest that selection of novel interventions may need to be tailored to each patient’s voluntary motor command “profile” rather than selected at the clinical population level. In the long term, if we can clearly identify different phenotypes and the neural mechanisms associated with each, then we can apply or develop specific interventions to target them. For example, Wolpaw and Thompson (2023) discuss the thought process for selecting different neural circuits to target with non-invasive stimulation paradigms designed to facilitate beneficial plasticity (e.g., operant reflex conditioning, paired associative stimulation). These paradigms have been applied mostly in the spinal cord injury population, and can initiate both local and widespread plasticity, ensuring that concurrent task-specific neurorehabilitation leads to beneficial rather than maladaptive plasticity. Our work is the first to indicate how the heterogeneity of the voluntary motor command in MS could be leveraged to guide the application of these targeted-plasticity paradigms.

### 4.6. Limitations

A main limitation of the study is that the approach to reverse engineer motor unit discharge patterns is only an indirect way to estimate neural inputs to spinal motoneurons, which of course cannot be measured directly in humans at present. Mitigating this limitation is the extent to which the reverse engineering paradigm has been developed on a foundation of direct measurements in animal preparations (e.g., Binder *et al*., 2020) and validated using realistic motoneuron models (e.g., Powers *et al*., 2012; Powers & Heckman, 2015, 2017; Johnson *et al*., 2017; Chardon *et al*., 2023). Additionally, there are methodological considerations that are common to other neurological populations, such as the potential contributions of changes in muscle fiber composition and medication use, and the unknown stability of our measurements over time in MS relative to controls.

This study produced a rich dataset that we will continue to explore fully. There are many open questions that are important for the interpretation and clinical relevance of our findings. For example, do our motor unit parameters relate to measures of motor and functional impairment in the MS group (e.g., Almuklass *et al*., 2018)? Are they associated with lesion locations, the extent of gray matter atrophy, etc.? How different would the findings be when testing the less-affected leg? Additionally, inclusion of quantitative testing of vibratory sensation, spasticity, and cognitive function would have enhanced our ability to interpret our motor unit findings.

## Data Availability

All data produced in the present study are available upon reasonable request to the authors.

## Author contributions

All experiments were performed at Washington University School of Medicine in St. Louis, MO.

Conceptualization: LM, KL, RN, and AC

Funding acquisition: LM, KL, RN, and AC

Methodology: LM, KL, JB, FN, RN, AC

Investigation: LM, SS

Data curation: LM, SS

Formal Analysis: LM, KL

Visualization: LM, KL

Writing – original draft: LM

Writing – review and editing: LM, KL, SS, JB, FN, RN, AC

## Acknowledgments

We thank the study participants for their time and effort, the John L. Trotter MS Center at Washington University for assistance with recruitment of MS patients, Tanner Reece, MS for assistance with data collection and preliminary analyses, and Daniel Free, PhD for assistance with data collection and code to automate tracking of motor units across trials.

## Supplemental Material

**Table S1.** Linear mixed effect model results for each reverse engineering variable: *Age, Sex, TQ_recrt_c*.

|  | Covariates |  |  |  |  |  |  |  |  |
| --- | --- | --- | --- | --- | --- | --- | --- | --- | --- |
|  | *Age* |  |  | *Sex* |  |  | *Torque at Recruitment* |  |  |
| | Bartlett | *p* | $\beta$<br>[CI] | Bartlett | *p* | mean diff<br>[CI] | Bartlett | *p* | $\beta$<br>[CI] |
| Delta-F | 20.5 | **< 0.0001** | -0.03<br>[-0.04, -0.02] | 2.1 | 0.15 | -0.28<br>[-0.67, 0.10] | 1.1 | 0.30 | 0.005<br>[-0.004, 0.014] |
| Norm delta-F | 16.2 | **< 0.0001** | -0.003<br>[-0.004, -0.001] | 0.98 | 0.32 | -0.02<br>[-0.05, 0.02] | 150.6 | **< 0.0001** | -0.009<br>[-0.010, -0.008] |
| Duration Ratio | 4.0 | **0.047** | -0.003<br>[-0.005, -0.00006] | 0.006 | 0.94 | 0.003<br>[-0.07, 0.08] | 1353.1 | **< 0.0001** | 0.022<br>[0.021, 0.023] |
| Braceheight | 1.3 | 0.26 | 0.0005<br>[-0.0003, 0.0013] | 0.05 | 0.82 | -0.003<br>[-0.03, 0.02] | 4.7 | **0.030** | 0.0012<br>[0.0002, 0.0022] |
| Acceleration amplitude | 9.3 | **0.002** | -0.015<br>[-0.024, -0.006] | 4.6 | **0.033** | -0.31<br>[-0.58, -0.03] | 48.9 | **< 0.0001** | -0.029<br>[-0.036, -0.021] |
| Acceleration slope | 0.23 | 0.63 | 0.002<br>[-0.005, 0.009] | 4.6 | **0.033** | -0.24<br>[-0.45, -0.03] | 105.7 | **< 0.0001** | 0.044<br>[0.036, 0.052] |
| Acceleration duration | 0.15 | 0.69 | -0.0004<br>[-0.002, 0.001] | 0.60 | 0.44 | 0.02<br>[-0.03, 0.07] | 812.9 | **< 0.0001** | -0.041<br>[-0.044, -0.038] |
| Rate attenuation slope | 0.47 | 0.49 | -0.0004<br>[-0.002, 0.0008] | 1.5 | 0.22 | -0.02<br>[-0.06, 0.01] | 90.6 | **< 0.0001** | 0.007<br>[0.005, 0.008] |
| Firing rate at recruitment | 5.1 | **0.024** | -0.014<br>[-0.025, -0.002] | 1.6 | 0.21 | -0.22<br>[-0.57, 0.13] | 143.7 | **< 0.0001** | 0.046<br>[0.038, 0.053] |
| Firing rate range | 12.6 | **0.0004** | -0.03<br>[-0.05, -0.02] | 4.4 | **0.035** | -0.57<br>[-1.08, -0.07] | 693.0 | **< 0.0001** | -0.106<br>[-0.113, -0.100] |
| Maximal firing rate | 5.4 | **0.020** | -0.03<br>[-0.05, -0.007] | 3.0 | 0.085 | -0.66<br>[-1.34, 0.03] | 528.5 | **< 0.0001** | -0.075<br>[-0.080, -0.069] |
| Firing rate at de-recruitment | 4.2 | **0.040** | 0.01<br>[0.001, 0.02] | 1.8 | 0.18 | -0.22<br>[-0.55, 0.10] | 148.3 | **< 0.0001** | 0.042<br>[0.035, 0.048] |
| Recruitment torque | 3.7 | 0.054 | -0.03<br>[-0.05, -0.0002] | 3.2 | 0.076 | -0.69<br>[-1.44, 0.05] | NA | NA | NA |
| De-recruitment torque | 0.25 | 0.62 | 0.01<br>[-0.03, 0.04] | 0.58 | 0.45 | 0.42<br>[-0.62, 1.45] | 2709.5 | **< 0.0001** | 0.40<br>[0.38, 0.41] |
| Duration | 0.40 | 0.53 | 0.01<br>[-0.03, 0.05] | 0.34 | 0.56 | -0.36<br>[-1.53, 0.83] | 9471.1 | **< 0.0001** | -0.79<br>[-0.81, -0.78] |
| Recruitment Range | 0.009 | 0.93 | 0.001<br>[-0.03, 0.03] | 0.06 | 0.81 | 0.11<br>[-0.82, 1.04] | 12.1 | **0.0005** | 0.014<br>[0.006, 0.022] |

